# Pan[dem]ic! Rational Risk Avoidance During a Health Pandemic

**DOI:** 10.1101/2021.05.28.21257983

**Authors:** Edward N. Okeke

## Abstract

During a health pandemic health workers have to balance two competing objectives: their own welfare vs. that of their patients. Intuitively, attending to sick patients during a pandemic poses risks to health workers because some of these patients could be infected. One way to reduce risk is by reducing contact with patients. These changes could be on the extensive margin, e.g., seeing fewer patients; or, more insidiously, on the intensive margin, by reducing the duration/intensity of contact. This paper studies risk avoidance behavior during the Covid-19 pandemic and examines implications for patient welfare. Using primary data on thousands of patient-provider interactions between January 2019 and October 2020 in Nigeria, I present evidence of risk compensation by health workers along the intensive margin. For example, the probability that a patient receives a physical examination has dropped by about a third. I find suggestive evidence of negative effects on health outcomes.

## 1 Introduction

The Covid-19 pandemic has changed the world as we know it. As of the time of writing, more than 130 million people worldwide have been infected, with three million dead (World Health Organization, 2021). The scale of the devastation can be hard to comprehend. Entire countries have been shut down in response (sometimes more than once), hospitals have been overrun (and continue to be in some regions), and millions have lost their jobs and livelihoods. Life expectancy, for the first time in decades, has reversed direction in some countries (Andrasfay and Goldman, 2021). Health workers, who have been on the frontlines of the pandemic, have been rightly celebrated as heroes.^1^ Even now, a year into the pandemic, they continue to operate in a high-stress high-uncertainty environment. The effects of this stress are now starting to emerge (Mehta et al., 2021; Rossi et al., 2020).

A key issue that has not attracted enough attention is that during this pandemic health workers have had to balance two objectives that have often been in conflict: their own welfare vs. that of their patients; on the one hand helping their patients and, on the other, protecting themselves. An illustrative anecdote that has circulated widely is of health workers abandoning their posts for fear of getting infected. While such behavior might be extreme, it helps to highlight the conflict that health workers face. By attending to sick patients during a pandemic, health workers face heightened risk of exposure (Kursumovic et al., 2020; Shah et al., 2020). This risk rises with case counts in the population, uncertainty about who is infected, and the amount of contact—frequency, duration and intensity—with sick patients.^2^ A rational health worker would try to reduce their exposure. Case counts are not under their control; uncertainty can be reduced by testing but has limited utility in the acute care setting because the results are not instantaneous,^3^ the one lever that is clearly within the control of the health worker is reducing contact with sick, and potentially infected, patients.^4^

Consistent with risk reduction, there are anecdotes of health workers refusing to provide care to patients during the pandemic. Risk avoidance may also be more subtle: for example, health workers might temporarily shut down their practice (for those who have this option), they might turn down risky assignments (e.g., nurses refusing to attend to Covid-19 patients), or might strategically call in sick.^5^ One can think of this as reducing contact with patients along the *extensive* margin. Such behavior would have implications for patient access to health care. More insidiously, health workers might reduce their contact with patients along the *intensive* margin: for example by spending less time with them or avoiding certain procedures that they believe carry more risk. This has implications for quality of patient care. The key issue here is that this type of risk avoidance behavior would potentially affect all patients since health workers cannot readily distinguish between who has the disease and who does not. Risk avoidance behavior, whether it is on the intensive or extensive margins, is therefore not only theoretically interesting, but has obvious policy relevance because of its potential impact on patient outcomes.

This paper examines risk avoidance behavior by health workers during the Covid-19 pandemic. The analysis primarily draws on data from patient surveys conducted in 288 primary health care health facilities in Nigeria over a two-year period between 2019 and 2020.^6^ I observe repeated cross-sections of patients treated in the same health facility and, in many cases, by the same health workers over time. I exploit the longitudinal nature of these data, using a range of fixed effects and difference-in-difference models, to draw fairly robust (and precise) conclusions about how health worker behavior has changed during the pandemic and how this has impacted patient care. In addition to these primary data, I will draw on surveys administered to health facility managers and health workers between August and October 2020 to shed light on how frontline health facilities reacted to the pandemic, and to examine health worker beliefs and perceptions.

I find evidence that health workers have mainly compensated on the intensive margin. Controlling for patient, facility and, in some cases, health worker characteristics, patients were 25% less likely to be asked recommended questions during the pandemic, they were 38% less likely to receive procedures that involved physical contact, including physical examinations, examinations with a stethoscope, and even blood pressure or temperature checks, and they were 35% less likely to receive supplementary health information from their provider. I obtain similar results using health facility fixed effects models (comparing patients with similar characteristics seen in the same health facility before and during the pandemic), health worker fixed effects models (comparing patients with similar characteristics seen by the same health worker before and during the pandemic), and difference-in-difference models (comparing the change in outcomes for patients seen in a given facility in the months before/after the start of the pandemic in 2020 to the same change in 2019). I show that these effects, while present for all patients, are larger for patients who presented with symptoms associated with Covid-19, providing further evidence that this is risk avoidance.

The concern is that this may impact patient health, adding to the toll of the pandemic. To examine this, I look at the delivery outcomes of women during the pandemic and, specifically, at the likelihood of postpartum complications. Looking at delivery outcomes is attractive from an econometric standpoint because women conceived long before the pandemic so the timing is unrelated to the pandemic, and unlike other more discretionary kinds of care, delivery cannot be put off until a more opportune time, drastically curtailing the role of selection. I have data on the delivery outcomes of 34,195 women randomly sampled from the communities served by the 288 primary health facilities. Drawing on data from in-home interviews conducted with these women approximately four months after their delivery, and using an event study design, I show that women who delivered after the start of the pandemic were significantly more likely to experience a postpartum complication compared to women who delivered just before. This increase is not driven by changes in women’s characteristics or by changes in healthcare utilization. I show that the results are robust to an alternative difference-in-difference empirical strategy that leverages data from women’s birth histories. There are other channels through which the pandemic might impact women’s delivery outcomes and so I do not claim that these effects are solely due to worse care but, at a minimum, they provide suggestive evidence that health worker responses to the pandemic may be negatively impacting health outcomes.

The results in this study have immediate policy relevance given that the Covid-19 pandemic is still ongoing. That the changes documented in this paper were evident more than seven months into the pandemic and showed no signs of leveling off, suggests that these are longstanding changes that might take time to reverse. Even though vaccines have become available, the role of mutated variants remains a wildcard. Vaccination may therefore not eliminate risk avoidance behavior. Additionally, based on current trends, it will likely take years for poorer countries to achieve anything close to full vaccination (Dyer, 2020). So what can policy makers do? In terms of policy solutions, increased monitoring of healthcare workers is unlikely to solve this problem, not only because it is hard to monitor some of these dimensions of care delivery, but also because it does not address the underlying factors that precipitate this behavior. A likely more effective solution is to compensate health workers for the increased risk that they face, e.g., by paying them a hazard allowance; plus greater investments in high-quality protective equipment, which is often lacking in resource-poor settings.^7^

This paper is the first to systematically examine risk avoidance behavior by health workers during the Covid-19 pandemic. It makes a novel and important contribution to the burgeoning pandemic literature. Prior work has shown that individuals adjusted their behavior in expected ways in response to the pandemic; for example, by reducing non-emergent healthcare utilization (Czeisler et al., 2020; Birkmeyer et al., 2020), but there has been little prior work looking at how the supply-side has responded to the pandemic. Additionally, while there has been some discussion in the literature of the secondary health effects of the pandemic (Green, 2020), thus far there has not been a lot of convincing empirical evidence. Beyond the pandemic-related literature, this paper makes a contribution to a broader economic literature on risk compensatory behavior (Dupas, 2011; Oster et al., 2013; Godlonton et al., 2016). This paper studies how health care workers respond to the higher (work-related) mortality risk during a contagious disease pandemic. It demonstrates how health workers acting in their individual best interests, in the best economic tradition, can lead to socially sub-optimal outcomes because of the externalities these actions impose on patients.

The rest of the paper proceeds as follows: Section 2 provides relevant institutional details, Section 3 describes the data, Section 4 describes the empirical strategy, results are presented in Section 5, and in Section 6 I discuss the findings. Section 7 concludes.

## 2 Background

### 2.1 Covid-19 in Nigeria

The Covid-19 epidemic was declared a pandemic by the World Health Organization on March 11, 2020. In March 2020, Nigeria had registered only a few cases of Covid-19, primarily among international travelers in the city of Lagos, a commercial center with an international airport. In response, international flights were suspended on March 23. By April there was evidence of community spread with cases confirmed in multiple states (Amzat et al., 2020). Stay-at-home orders were imposed around the country in April. Essential workers, such as health workers and security personnel, were exempt. These orders began to be lifted in May, and were replaced by less restrictive night-time curfews. By September/October 2020, all restrictions had largely been lifted and life in Nigeria had essentially returned to normal. International flights resumed in September 2020. By this point, only a few hundred daily cases were being reported nationwide (see Figure 1). A second wave appears to have begun in December 2020. My data cover the entirety of the first wave up to November 2020. Based on the trajectory in Nigeria, and illustrated by Figure 1, April is used as the start month of the pandemic.

**Figure 1:**
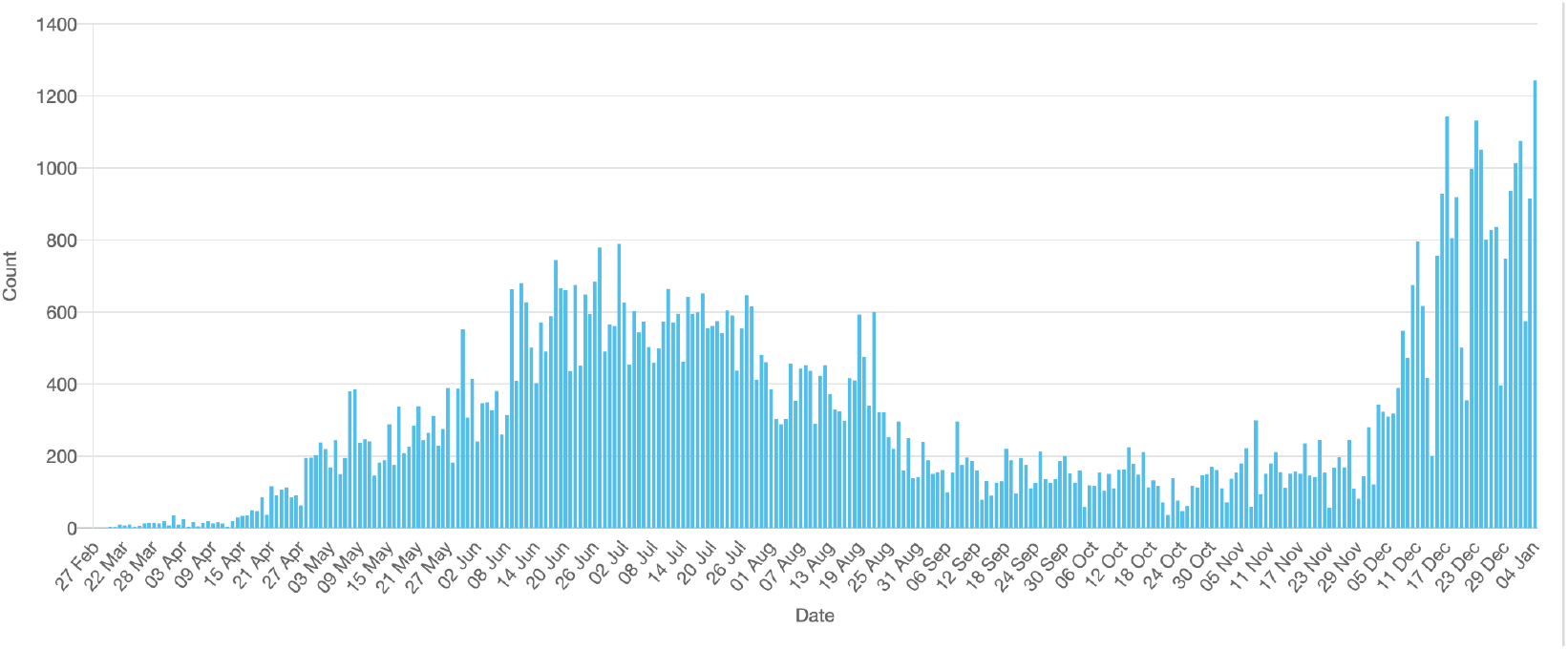
Daily Counts of Positive Covid-19 Tests in Nigeria (March 2020-Jan 2021) Note: Figure 1 shows daily counts of positive Covid-19 tests in Nigeria. Figure is from the Nigeria Centre for Disease Control (NCDC) website. Available at ncdc.gov.ng. Accessed January 8, 2021

As of March 14 2021, Nigeria had registered 160,657 total cases of Covid-19 cumulatively, and 2,013 deaths, among a population of more than 200 million, though this is almost certainly an undercount (Nigeria Centre for Disease Control (NCDC), 2021).

### 2.2 Study design and participants

This study uses data from a large research study conducted in 288 public primary health care facilities distributed across four states.^8^ For context, Nigeria operates a tiered health care system with primary health centers forming the base of the pyramid (general hospitals act as referral centers and form the middle of the pyramid, and University Teaching hospitals are at the apex of the pyramid, providing the most advanced care). Primary health facilities provide a broad range of preventive, outpatient, and inpatient services and are the point of entry for most patients into the health care system. The health care facilities were selected with the help of government health officials. The list was finalized in the fall of 2018. The selected facilities represent approximately 12% of available primary health facilities (including smaller health posts and dispensaries which provide only a limited range of services). They should be considered broadly representative. Combined, they provide health care services to more than one million individuals. These health facilities are the main source of care for households living in the surrounding communities (these communities form what is known as the catchment or service area). Table 1 provides a description of these facilities.

**Table 1:**
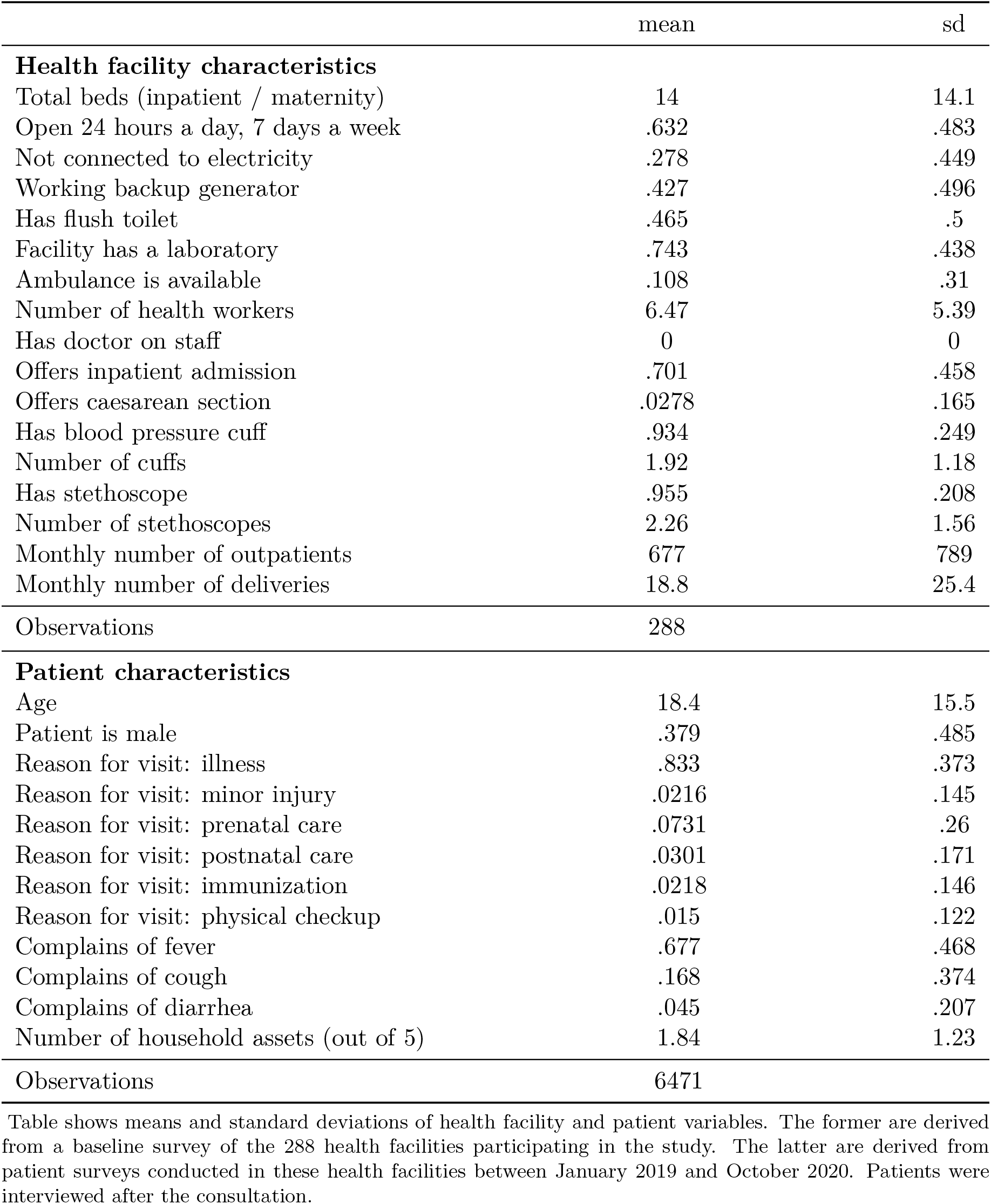
Summary Statistics: Health Facilities and Patients

|  | mean | sd |
| --- | --- | --- |
| **Health facility characteristics** |  |  |
| Total beds (inpatient / maternity) | 14 | 14.1 |
| Open 24 hours a day, 7 days a week | .632 | .483 |
| Not connected to electricity | .278 | .449 |
| Working backup generator | .427 | .496 |
| Has flush toilet | .465 | .5 |
| Facility has a laboratory | .743 | .438 |
| Ambulance is available | .108 | .31 |
| Number of health workers | 6.47 | 5.39 |
| Has doctor on staff | 0 | 0 |
| Offers inpatient admission | .701 | .458 |
| Offers caesarean section | .0278 | .165 |
| Has blood pressure cuff | .934 | .249 |
| Number of cuffs | 1.92 | 1.18 |
| Has stethoscope | .955 | .208 |
| Number of stethoscopes | 2.26 | 1.56 |
| Monthly number of outpatients | 677 | 789 |
| Monthly number of deliveries | 18.8 | 25.4 |
| Observations | 288 |  |
| **Patient characteristics** |  |  |
| Age | 18.4 | 15.5 |
| Patient is male | .379 | .485 |
| Reason for visit: illness | .833 | .373 |
| Reason for visit: minor injury | .0216 | .145 |
| Reason for visit: prenatal care | .0731 | .26 |
| Reason for visit: postnatal care | .0301 | .171 |
| Reason for visit: immunization | .0218 | .146 |
| Reason for visit: physical checkup | .015 | .122 |
| Complains of fever | .677 | .468 |
| Complains of cough | .168 | .374 |
| Complains of diarrhea | .045 | .207 |
| Number of household assets (out of 5) | 1.84 | 1.23 |
| Observations | 6471 |  |

Table shows means and standard deviations of health facility and patient variables. The former are derived from a baseline survey of the 288 health facilities participating in the study. The latter are derived from patient surveys conducted in these health facilities between January 2019 and October 2020. Patients were interviewed after the consultation.

Some of the health facilities took part in an intervention in which they received small grants designated for quality improvement. One-third of the health facilities were randomly selected to receive this grant. All of the analytical models include either facility or health worker fixed effects, which hold fixed any facility-level factors, including any interventions provided. I will also test whether there are heterogeneities in treatment effect by intervention arm (there are not).

## 3 Data

Baseline data collection took place in all 288 health facilities between December 2018 and March 2019. During this visit, we collected data about the characteristics of the health facility including the number of beds, number of health providers, and structural characteristics such as source of power and availability of running water. We also administered brief surveys to all health workers that were available in the health facility on the day of the visit. Lastly, we interviewed patients who visited the health facility on that day for care. These patients were interviewed after they had received care.^9^ The patients (or their caregivers) were asked questions about the care they received. For example, they were asked whether they were physically examined by the health provider, whether their blood pressure and temperature was measured during the visit, and whether a stethoscope was used at any point during the consultation.

For each patient we also collected data on their characteristics: their age, gender, and the reason for the visit (e.g., illness, treatment for minor injury, prenatal care). For sick patients, I have data on their presenting complaints (e.g., fever, cough, headache). I therefore have much of the same data available to the health provider. In addition, we collected some socioeconomic data: ownership of five specified household assets. These data allow me to credibly control for differences in patient composition over time. Patient descriptives are also reported in Table 1. All interviews were conducted by trained research assistants, employed by a local University partner, using survey software on a computer tablet.

We returned to these facilities three more times between 2019 and 2020. The second visit was conducted between August and November 2019. The third visit was between January and April 2020. The fourth visit was between August and October 2020. During each of these visits, we surveyed patients again using the same protocols as the baseline visit. I therefore observe a repeated cross-section of patients receiving care in each of the 288 health facilities between January 2019 and November 2020. The aggregate number of patients interviewed in each month is shown in Table A.1. The in-charge of the facility (the facility manager) and health providers were surveyed again during Visit 4 (August-October 2020). A new Covid-19 module was added to both questionnaires for Visit 4. The former collected information about changes made in the health facility in response to the pandemic, while the latter collected information about health worker perceptions and beliefs about the SARS-CoV-2 virus.

In addition to these visits described above we also carried out multiple unannounced visits to each of these health facilities (these will sometimes be referred to as audit visits). During these visits the research assistant recorded whether the facility was open and verified whether each health worker employed in the facility was present (the staff list was compiled from the staff register at baseline and periodically updated). For each individual health worker *i*, I therefore observe whether they were present during visit *v*. As part of the audit visit, the research assistant also counted, and recorded, the number of patients seen waiting in the waiting area. This gives us a measure of patient volume. We tried to make an unscheduled visit to each health facility at least once per calendar trimester. The median facility received three visits between June 2019 and October 2020 (mean = 3.3). Together with the data from the baseline visit, which was not pre-scheduled, I observe, objectively verified, health worker availability in the health facility between January 2019 and October 2020. The aggregate number of health workers observed in each month is shown in Table A.2.

To summarize, there are four primary sources of data, obtained from health facilities, that will be used in the analysis: (1) Health facility surveys (administered to the in-charge during Visit 4), (2) Health worker surveys (administered to all health workers present during Visit 4), (3) Patient surveys (administered to patients in each health facility during Visits 1-4), and (4) Audit visits (conducted roughly once per trimester). For clarity the timeline of visits and the data collected during each visit are laid out visually in Table 2.

**Table 2:**
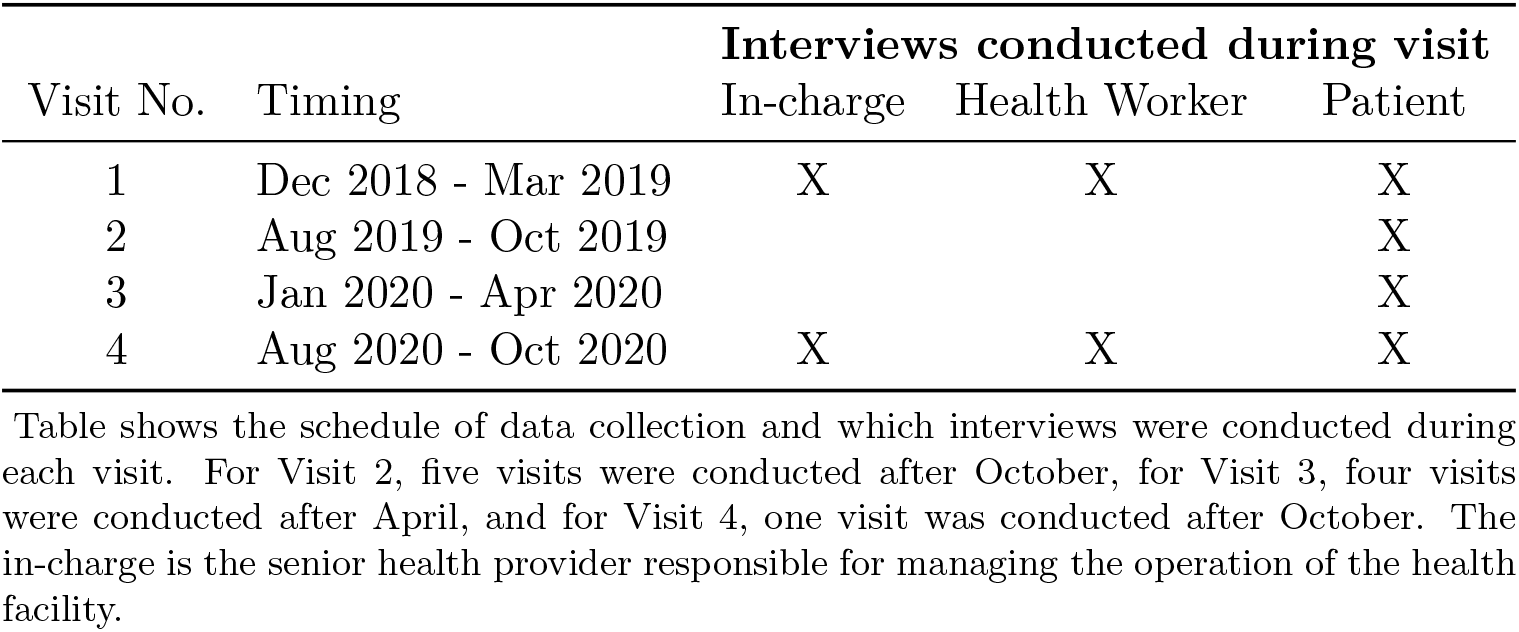
Schedule of data collection

| Visit No. | Timing | **Interviews conducted during visit** |  |  |
| --- | --- | --- | --- | --- |
|  |  | In-charge | Health Worker | Patient |
| 1 | Dec 2018 - Mar 2019 | X | X | X |
| 2 | Aug 2019 - Oct 2019 |  |  | X |
| 3 | Jan 2020 - Apr 2020 |  |  | X |
| 4 | Aug 2020 - Oct 2020 | X | X | X |

Table shows the schedule of data collection and which interviews were conducted during each visit. For Visit 2, five visits were conducted after October, for Visit 3, four visits were conducted after April, and for Visit 4, one visit was conducted after October. The in-charge is the senior health provider responsible for managing the operation of the health facility.

### 3.1 Data on delivery outcomes

For the research project we randomly sampled approximately 120 pregnant women from the service area of each participating primary health facility. To sample women, we first randomly selected a community from the service area and then conducted a door-to-door census to identify all pregnant women in the community.^10^ All women who gave consent were enrolled (pregnancy was the only eligibility criterion). The refusal rate was less than 0.1%. This process was repeated—randomly selecting another community, conducting a census, and enrolling all women—until the desired target number of approximately 120 women was reached. Recruitment and enrollment took place between July 2019 and April 2020.

These women participated in some study interventions. Approximately half were randomly offered modest cash incentives conditioned on use of pregnancy and delivery care. The incentive was announced at baseline but payment was made later, after the delivery. This was crosscut with another intervention that provided information, at baseline, about pregnancy risk factors. These interventions were randomly assigned at the service area-level. This paper is not about these interventions but, to control for their effects, I include indicators for any interventions provided in the community. Some models also include service area or community fixed effects, which hold any area-level factors fixed including any interventions provided. I will also test for heterogeneity in the effect of the pandemic across the different intervention groups.

On enrollment, each participating woman completed an interview during which we collected basic demographic information and a full retrospective birth history (i.e., all births prior to enrollment). For the most recent delivery within the last three years (since January 2016) we collected information about some postpartum (delivery) complications. Each woman completed another in-home follow-up interview approximately four months after delivery (of the child that was in utero at enrollment). This interview collected detailed information about health care utilization during the current pregnancy, and major postpartum complications. All interviews were conducted by trained data collectors. Ethical approval for the research project was granted by RAND’s Human Subjects Protection Committee and the Ethics Committee of Aminu Kano Teaching Hospital, Kano.

#### Descriptives

36,607 pregnant women from 1,365 communities were enrolled into the study at baseline. 34,299 gave birth between July 2019 and December 2020, 28 died while pregnant, 1,841 had an early pregnancy termination, 190 were never pregnant, 177 were not located at follow-up, and 72 did not give consent for the follow-up. Table 3 provides baseline summary statistics.

**Table 3:** Baseline Summary Statistics of Enrolled Women

|  | mean | sd |
| --- | --- | --- |
| Age at enrollment | 25.1 | 6.24 |
| Moslem religion | .734 | .442 |
| Hausa or Fulani ethnicity | .703 | .457 |
| No schooling | .289 | .453 |
| Islamic schooling | .331 | .47 |
| Some formal schooling | .381 | .486 |
| Number of prior births | 2.64 | 1.5 |
| First child | .332 | .471 |
| Age is 15 years or younger | .0135 | .116 |
| Age is 35 years or older | .101 | .301 |
| Polygamous household | .19 | .392 |
| Number of household assets (out of 10) | 1.86 | 1.85 |
| Observations | 36607 |  |

Table shows means and standard deviations of variables derived from a baseline survey of women enrolled in the study. The women were randomly sampled from communities served by the 288 participating health facilities.

## 4 Empirical Strategy

The analysis will answer the following questions: First, how did health facilities react to the pandemic? What adaptive changes did they make in response? Second, how well-informed were health workers about the SARS-C0V-2 virus? Third, how did the pandemic affect health worker behavior? What impacts, if any, did this have on patient care? The last part of the analysis examines women’s delivery outcomes during the pandemic to shed light on health implications. For the first two questions, the analysis will primarily be descriptive. The analysis will draw on the Covid-19 module added to the health facility and health worker surveys administered during Visit 4 (August-October 2020). Below I discuss how I intend to approach answering the other questions.

### 4.1 How did the pandemic affect health worker behavior?

There are two main outcomes that I will examine: (1) health worker availability during the pandemic, and (2) the ‘quality’ of patient interactions. I begin with health worker availability. Is there any evidence that this reduced during the pandemic as has been (anecdotally) reported? Increased absence from work during the pandemic would be suggestive of an extensive margin response. To answer this question, I propose to estimate the following linear probability model:

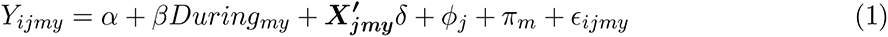

*Y_ijmy_* denotes whether health worker *i* in health facility *j* was present during an unscheduled facility visit in month *m* and year *y*. *During_my_* is equal to one if the visit occurred after the start of the pandemic, i.e. between April and November 2020, and zero otherwise. The model includes calendar month fixed effects (*π_m_*), to control for any seasonal trends in health worker availability, and facility fixed effects (*φ_j_*) to control for fixed facility-level factors. *X^t^δ* denotes time-varying controls. I control for the day of the week of the visit (health workers might be less likely to be present on certain days, e.g., Fridays), and the time of day (health workers might be less likely to be present later in the day). *β* is the parameter of interest. The standard errors are clustered at the facility level to account for the repeated observations within facility. This basic specification compares a given facility, visited in month *m* during the pandemic, to itself in the same month in 2019. I will also estimate an augmented specification that includes health worker fixed effects (*α_i_*). This is a more demanding specification that compares the same health worker to themselves. I lose some observations because not all health workers were present throughout the period.

One concern with this strategy, potentially, is that there could be pre-existing temporal trends that are captured in *β*. Though it is not obvious that health worker availability in the facility should exhibit such trends beyond within-year seasonality, which is already accounted for in the model with the inclusion of calendar month fixed effects, but to account for this I implement an alternative double-difference strategy. I first c ompare t he change in the outcome during the pandemic (April-November) to the months before the pandemic (January-March) in 2020 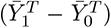 and take the same difference in 2019 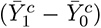. *Ȳ* denotes means. I then take the difference between these two estimates 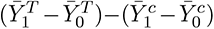. This double-difference gives us an unbiased estimate of the effect of the pandemic that is uncontaminated by any pre-existing trends, provided that trends do not vary systematically by season. These trends are captured by 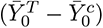, which can be estimated directly in the model. The model is parameterized as shown below:

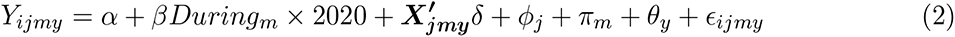

*During_m_* is equal to one if the visit occurred after March in either year, and zero otherwise. *φ_j_*, *π_m_*, and *θ_y_* are a full set of health facility, calendar month, and year fixed effects. *β* is the double-difference estimate. Note that Equation 2 does not include *During_m_* because the model includes calendar month fixed effects (*During_m_* is subsumed by *π_m_*).

#### 4.1.1 Quality of patient interactions

To test for changes in the quality of patient interactions, I will use a similar empirical strategy to that outlined above. The analysis uses the multiple waves of patient surveys. Because I observe patients receiving care in the same health facilities over time, I can estimate a facility fixed effects specification as in Equation 1, I can also estimate a health worker fixed effects specification, but with an important caveat: I only know the treating provider after Visit 1, i.e., from Visit 2 to Visit 4. Visit 1 was used to compile the staff register, which was then used to identify treating providers during later rounds. I also lose some observations where the health worker is only present in one wave. Because of this, the sample using the health worker fixed effects specification is substantially smaller, and different, from the other samples. For this reason I report the results in the Appendix, but I will show that all the results carry through. I will also estimate the double difference specification shown in Equation 2.

I have rich data on the quality of provider-patient interactions including: (i) whether the provider asked history-taking questions recommended by clinical guidelines, (ii) whether the provider examined the patient (literally, whether they touched the patient), (iii) whether the patient’s temperature was taken, (iv) whether the patient’s blood pressure was measured, and (v) whether a stethoscope was used at any point during the interaction. I also have data on more qualitative aspects of the interaction, e.g., whether the treating provider explained the diagnosis to the patient, whether they provided any health education related to the diagnosis, and whether they told the patient when to return for a follow-up visit. One can think of these as measuring the quality of the health provider’s communication with the patient.

At this point, one might be wondering whether patients are able to reliably report on these measures. As noted before patients were interviewed soon after the interaction (so events would still be fresh in their minds). To show that patient reports are in fact reliable, in the Appendix I compare patient reports to the report of an independent observer. A subset of clinical interactions were independently observed by a member of the research team who sat in a corner of the room where they could observe the interaction but did not otherwise say or do anything to interfere with the encounter. They recorded their observations on a structured form that collected some of the same information that was later asked of patients. These independent observations were conducted during Visit 3. So, for this subset, I have patient reports and the reports of an independent observer for the same encounter. I compare agreement between these two reports in Table A.3. I report the percentage agreement between the patients’ responses and the observers’ reports and Cohen’s kappa, a measure of interrater agreement. One does not expect perfect agreement, but we would hope to see substantial agreement. Percent agreement is generally high, and the kappa statistic indicates that there is substantial agreement between both reports.^11^

### 4.2 Health implications

Depending on how health workers responded to the pandemic, one can easily imagine downstream health effects. For example, if health workers were less likely to be available to provide care, this would affect access. Similarly, if the quality of patient interactions changed in important ways, this could also lead to worse outcomes for patients. To examine health implications I look at women’s delivery outcomes. Specifically, I examine the likelihood of experiencing postpartum complications. Looking at delivery outcomes is attractive for several reasons: first, women giving birth during the pandemic—at least over the period covered by my data—conceived long before the pandemic started, so there is no chance that pregnancy decisions were influenced by the pandemic; second, while some kinds of care can be deferred—and there is evidence that the pandemic influenced individuals to postpone care— see for example Czeisler et al. (2020) and Birkmeyer et al. (2020)—labor and delivery cannot be deferred. Both of these suggest that selection should not be an important concern, and women delivering just before the start of the pandemic should be similar to women delivering just after. The exogenous start of the pandemic therefore creates something akin to a local experiment. I will present evidence consistent with this later.

Why postpartum complications? Because one can potentially draw a line from this to the process (and quality) of care around delivery. I focus on two specific complications: infections and severe bleeding, both of which account for about 40% of all maternal deaths (Say et al., 2014).^12^ Not only are they substantively important, we also have good evidence that they are largely preventable. Good quality care reduces both the risk of severe bleeding (Begley et al., 2019; Gülmezoglu et al., 2009) and postpartum infections (World Health Organization, 2016). Therefore, if the quality of interactions between health workers and patients declined during the pandemic, one might expect to see an increase in delivery complications. Anecdotes of women being left to labor alone, because health workers were scared of getting infected, have circulated during the pandemic. Under such circumstances one can imagine early danger signs being missed. It is important to stress that this would not only affect women who delivered in the health facility but also women who delivered at home (or elsewhere), who might be taken to these facilities for emergency care. There should not be significant overlap between these conditions and Covid-19 but I will discuss interpretation more in Section 6.

The analysis uses data from women’s follow-up interviews (as a reminder, these interviews were conducted about four months after the delivery). A limitation of these data is that outcomes are self-reported but, given the relatively short recall period, I am cautiously optimistic that women are able to reliably recall these events. There is some evidence from validation studies that women can reliably recall delivery complications (Stewart and Festin, 1995; Sou et al., 2006), and similar outcomes have been used in prior work (Okeke et al., 2020; Díaz and Saldarriaga, 2019). To test for changes I use an event study design. This compares the outcomes of women delivering in the months just before vs. just after the start of the pandemic. As discussed earlier, the timing of conception is exogenous to the pandemic, as is the timing of labor (<1% of deliveries are by caesarean section), so there should be little difference in the characteristics of women delivering in the months before and after the start of the pandemic. I will present evidence of this.

The event study specification is shown in Equation 3 below.

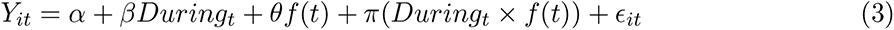

*Y_it_* denotes whether woman *i* who gave birth at calendar time *t* experienced a postpartum complication. *f* (*t*) is a linear function of time (which seems to fit the data reasonably well as I will show later). Time is measured in months and centered so that zero is the start month of the pandemic. *During_t_* is equal to 1 for *t ≥* 0. I allow the regression function to differ on both sides of the cutoff (*During_t_ × f* (*t*)). *β* is the coefficient of interest. The base specification compares deliveries occurring within eight months of the start of the pandemic, i.e., *t ∈* [*−*8, 8]. In covariate-adjusted specifications, I control for the mother’s age, level of schooling, ethnicity, religion and household wealth. I also control for any interventions provided. If women delivering just before and after the start of the pandemic have similar characteristics, an assumption I will validate later, including covariates should not alter the point estimates. The standard errors are adjusted for clustering among women in the same community. I will also report, for comparison, standard errors clustered at the service area level. They are similar.

I will present results from multiple robustness checks including: specifying time as a quadratic, including area fixed effects, and progressively shrinking the comparison time window from eight, to six, and then four months. The tradeoff is that, as I shrink the window, I lose observations and power. Finally, as an alternative empirical strategy, I will estimate a standard difference-in-difference model similar to Equation 2. This leverages the fact that I have data on postpartum complications for women’s last delivery prior to enrollment (from their retrospective birth history). For their most recent delivery within the three years preceding enrollment I know whether they experienced postpartum bleeding.^13^ For this outcome I can compare changes between January-March and April-November in 2020 (first difference) to changes between January-March and April-November in prior years (second difference). The model includes community, calendar year, and calendar month fixed effects. Standard errors are clustered at the community level.

## 5 Results

Figure 2 describes the adjustments made by primary health facilities. They implemented a range of changes that appear aimed at reducing the risk of transmission and infection in the facility. Some of these changes were inward-facing focusing on facility processes; these include more frequent cleaning of the facility and mask wearing by providers. Others were outward-facing and targeted towards patients, including social distancing requirements and screening for Covid-19 symptoms. Facility-focused measures were much more common than patient-focused measures: for example, mask wearing by providers, more frequent hand washing, and use of hand sanitizer were almost universally adopted.

**Figure 2:**
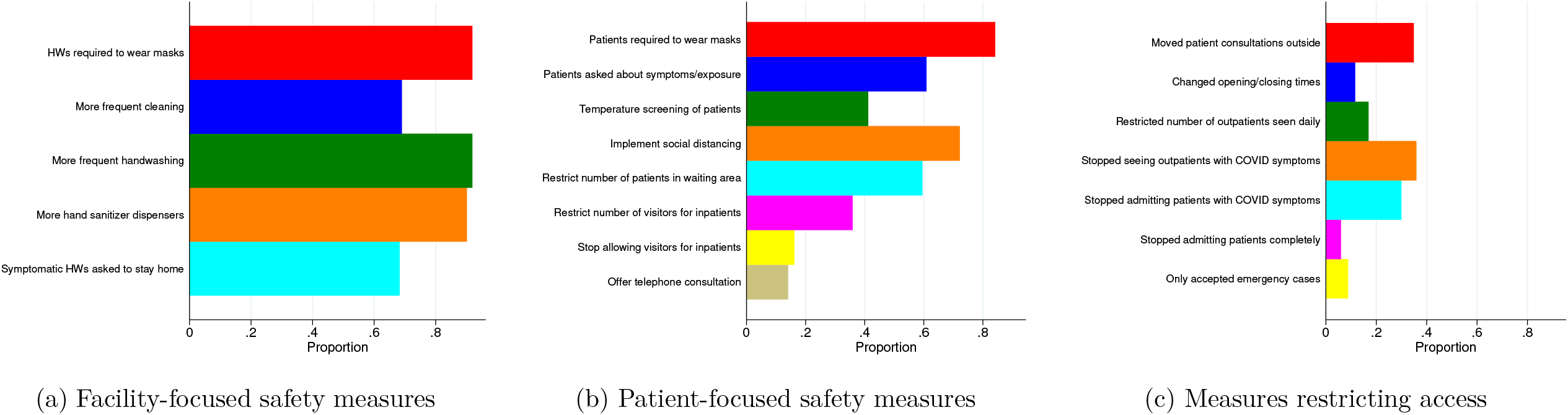
Health facility responses to the Covid-19 pandemic Note: The data are from a survey of 288 primary health facilities in Nigeria that was conducted between August and October 2020. HW denotes Health Worker.

The most common measure targeted towards patients was requiring masks/face coverings. However, this was not universal: about 15% of facilities reported not requiring patients to wear masks or face coverings. The next most frequently implemented patient measure was social distancing; implemented by about three-quarters of facilities. Only about 60% of facilities reported screening patients for symptoms and prior exposure to Covid-19, and about 40% reported implementing regular temperature screening. Since most facilities used manual mercury thermometers, checking temperature of all patients, and accompanying caregivers, may simply not have been feasible; plus it would have required fairly close contact with individuals which, as I will show later, health workers preferred to avoid.

In addition to these measures, which were largely expected, health facilities also adopted other measures that were more inimical. Some of these are particularly noteworthy: for example, 35% of health facilities reported moving patient consultations outside, a similar proportion of facilities stopped providing outpatient care to any individuals reporting Covid-19 symptoms, and 30% of facilities stopped admitting any patients with Covid-19 symptoms (a small percentage stopped admitting any patients at all). These changes are consistent with facilities broadly restricting access to individuals that *might* be infected with the virus. An undesirable side effect of these policies, however, is that they reduced patient access to care.

### 5.1 Health workers perceptions and beliefs

The data include responses from 1,324 health workers surveyed during Visit 4,^14^ and provide valuable insight into health workers’ perceptions and beliefs. There is both good news and bad. The good news is that nearly all health workers reported that they believed that the SARS-CoV-2 virus was real: 99% believed that it was definitely real and 1% believed that it was likely real. The bad news is that health workers appeared to systematically overestimate mortality associated with the SARS-CoV-2 virus. This is important because beliefs about lethality influence perceptions of risk. From the health worker’s perspective, the risk is obviously much greater if the virus has a 50% mortality rate compared to if the mortality rate is 0.1%. To assess beliefs about mortality associated with Covid-19, we posed a hypothetical, asking them how many people (out of 100) they thought would die if all 100 got infected with the virus. Their responses are summarized in Figure 3. Because of heaping, the responses are grouped.

**Figure 3:**
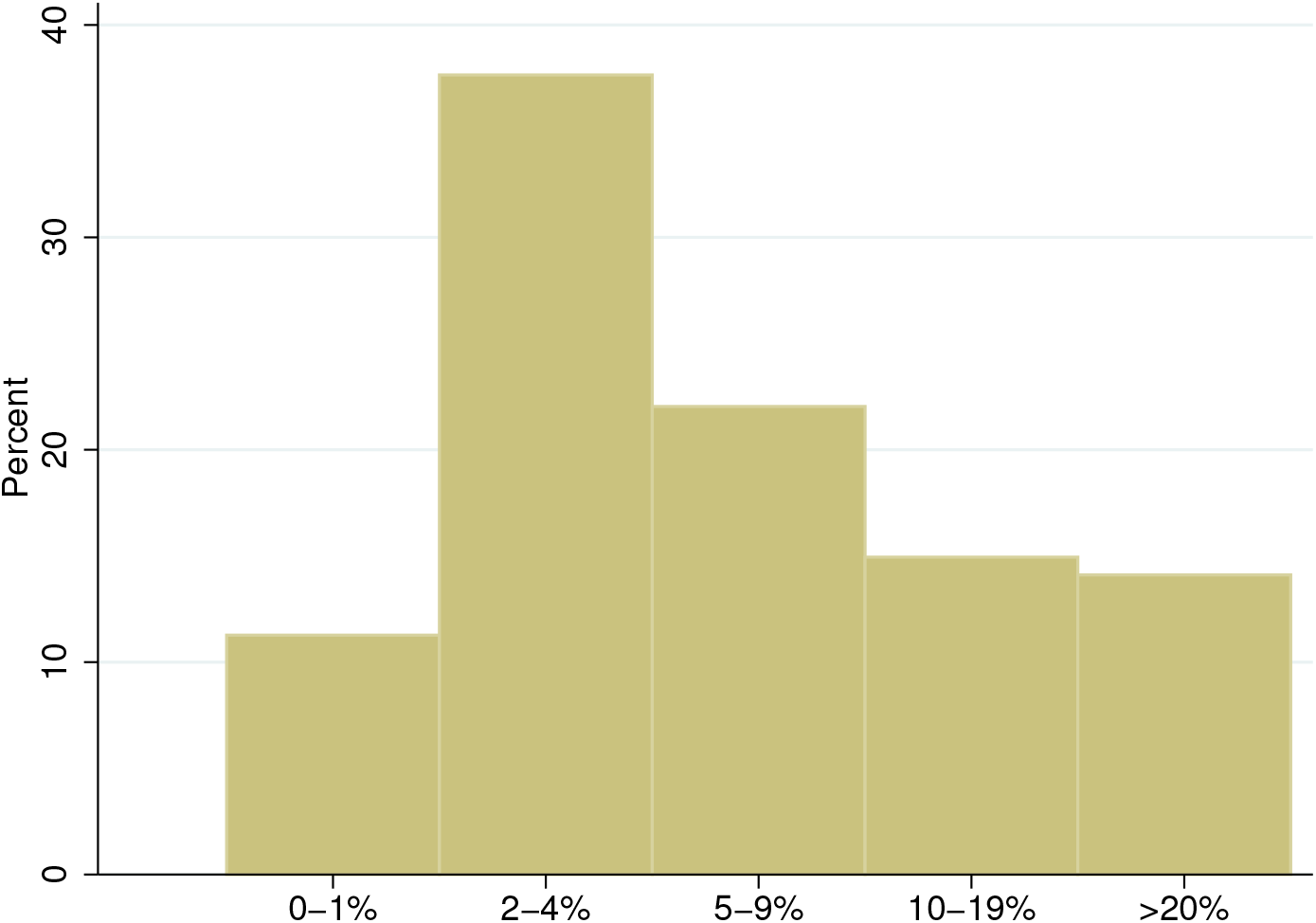
Health worker perceptions of mortality risk associated with Covid-19 Note: To assess perceptions health workers were asked how many people (out of 100) they thought would die if all 100 got infected with the SARS-CoV-2 virus. The data are from a survey of health workers in 288 primary health facilities in Nigeria that was conducted between August and October 2020.

The median/mean reported probability was 5%/8%.^15^ For context, the true probability is less than 1% (Ioannidis, 2021). In other words, the median health worker overestimated Covid-19 case fatality by more than 500%. Almost 1 in 3 workers (29%) believed that the fatality rate exceeded 10%, and about 1 in 7 (14%) believed that it exceeded 20%.^16^

For additional context these surveys were administered between August-November 2020 and not during the early months of the pandemic when there was still considerable uncertainty about case fatality. Overestimating mortality associated with Covid-19 is neither particularly unusual nor revelatory (Rothwell and Desai, 2020), what makes it important is the context. Like other individuals, health workers’ beliefs will influence their behavior. The key issue here are the consequences and impacts of these behaviors.

Figure 4 examines beliefs about how the virus is transmitted. For each mode of transmission health workers could choose one of five options: strongly agree, agree, uncertain, disagree, and strongly disagree. The results here are mixed. While, overall, health workers seemed reasonably well-informed—99% correctly stated, for example, that it was spread by coughing/sneezing—a non-trivial fraction harbored erroneous beliefs: 36% of health workers, for example, believed that the virus was transmitted through blood; a further 11% were uncertain. This becomes important in the context of providing care to patients. If a health worker erroneously believes that the virus can be transmitted by coming into contact with the blood of an infected patient, or is unsure, one can imagine that they would be extra-cautious about their contact with patients. 20% of workers also believed that it could be spread through sexual intercourse; 10% were uncertain. This is arguably less relevant for the health care setting.

**Figure 4:**
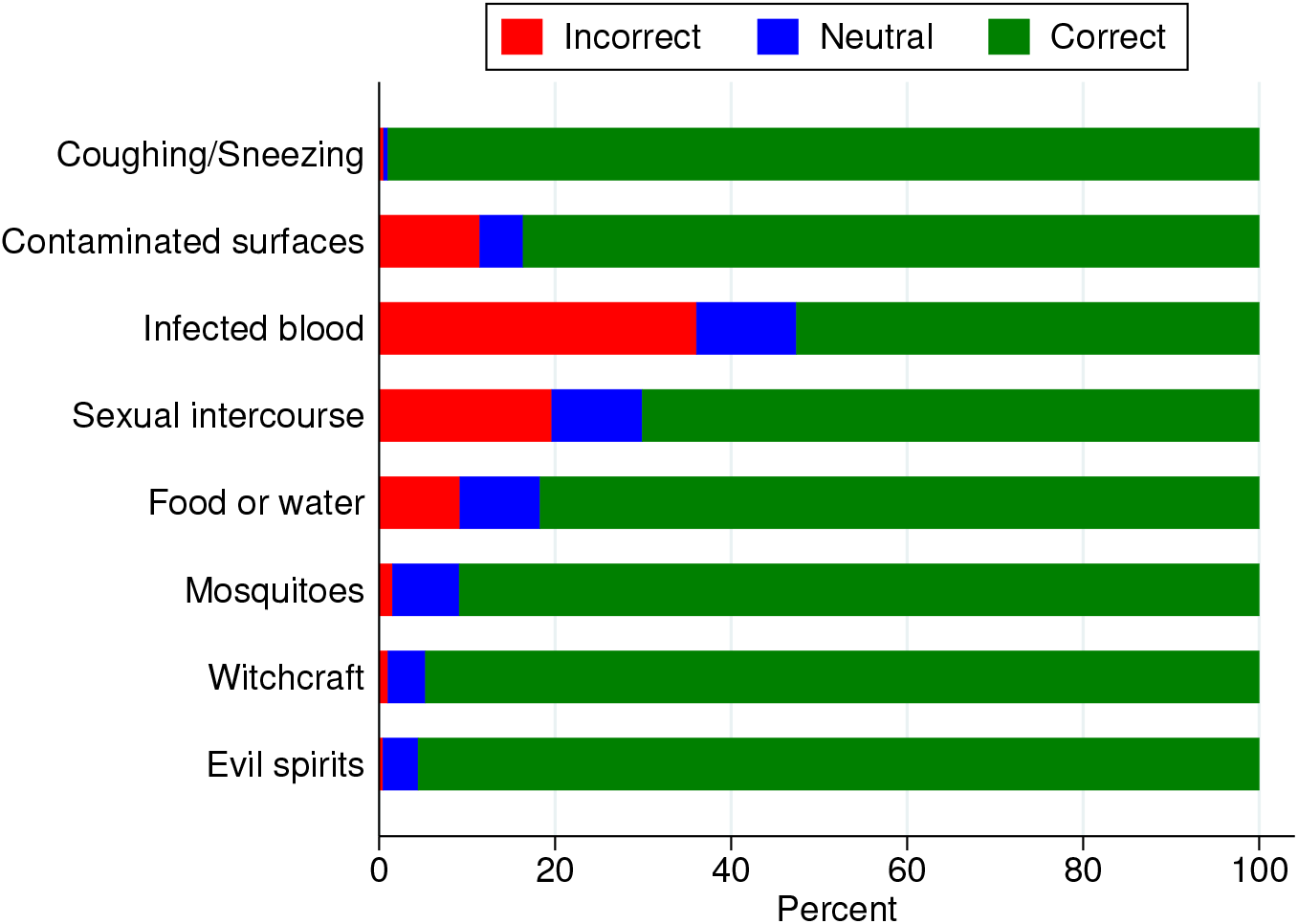
Health worker beliefs about how Covid-19 virus is transmitted Note: For each mode of transmission health workers could choose one of five options: strongly agree, agree, uncertain, disagree, and strongly disagree. Agreement (agree or strongly agree) is coded as correct for the first two modes of transmission (from top) and as incorrect for the other modes of transmission. Uncertain is coded as neutral. The data are from a survey of health workers in 288 primary health facilities in Nigeria that was conducted between August and October 2020.

Table A.5 examines whether worker characteristics are predictive of beliefs. The explanatory variables include age, sex, years of work experience, experience squared, and college-level education (at least a Bachelor’s degree or equivalent).^17^ The dependent variables are perceived mortality risk associated with Covid-19, worry about getting infected, and correct beliefs about transmission (I assign a score of +1 for correct responses, -1 for incorrect responses, and 0 for neutral responses and take the sum). Since these are count variables, I use a Poisson regression model. Average marginal effects are reported. The one variable that consistently predicts beliefs/perceptions is sex: male health workers reported higher perceived mortality rates for Covid-19, they were significantly more worried about getting infected, and had more incorrect beliefs about how SARS-CoV-2 is transmitted. There is some evidence that more highly educated health workers (those with at least a Bachelor’s degree) were more likely to hold accurate beliefs. They reported lower mortality, were less worried about getting infected, and were more likely to harbor correct beliefs about transmission, though this latter result is not statistically significant.

### 5.2 Effect on health worker behavior

I start by examining whether health workers were less likely to be present in the health facility during the pandemic. As anecdotal reports have suggested, were they less likely to show up to work? This would be consistent with risk avoidance along the extensive margin. I find no evidence of this in Figure 5. The probability that a health worker was present in the facility decreased by a statistically insignificant 3.1 percentage points; from 68.8% between January-March 2020, to 65.7% between April-November (P = 0.20), a relative decrease of 4.5%. In 2019, over the same period, it increased by 3.2 percentage points (*P* = 0.26). The corresponding regression results in Table 4 confirm this. Across all specifications, there is no statistically significant evidence that health workers were less likely to be present during the pandemic. It seems, therefore, that we can reject the narrative of the absconding health worker, at least in this setting.

**Figure 5:**
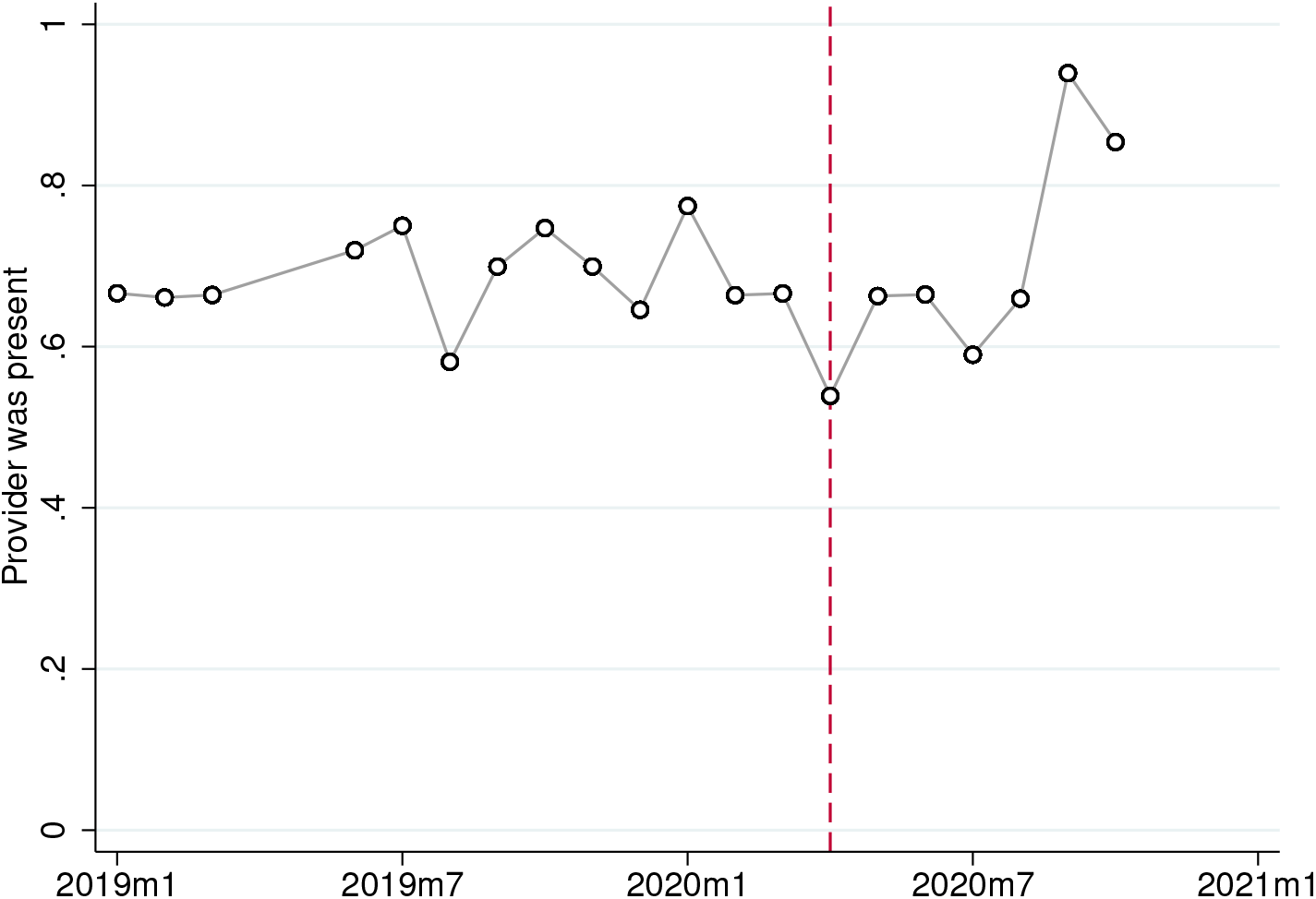
Probability that the health worker was present in the facility Note: Figure plots the probability that a health worker was present in the facility during an unscheduled visit. Data are averaged for each month. The number of health worker observations per month is shown in Table A.2. Dashed line denotes the start month of the pandemic in Nigeria.

**Table 4:**
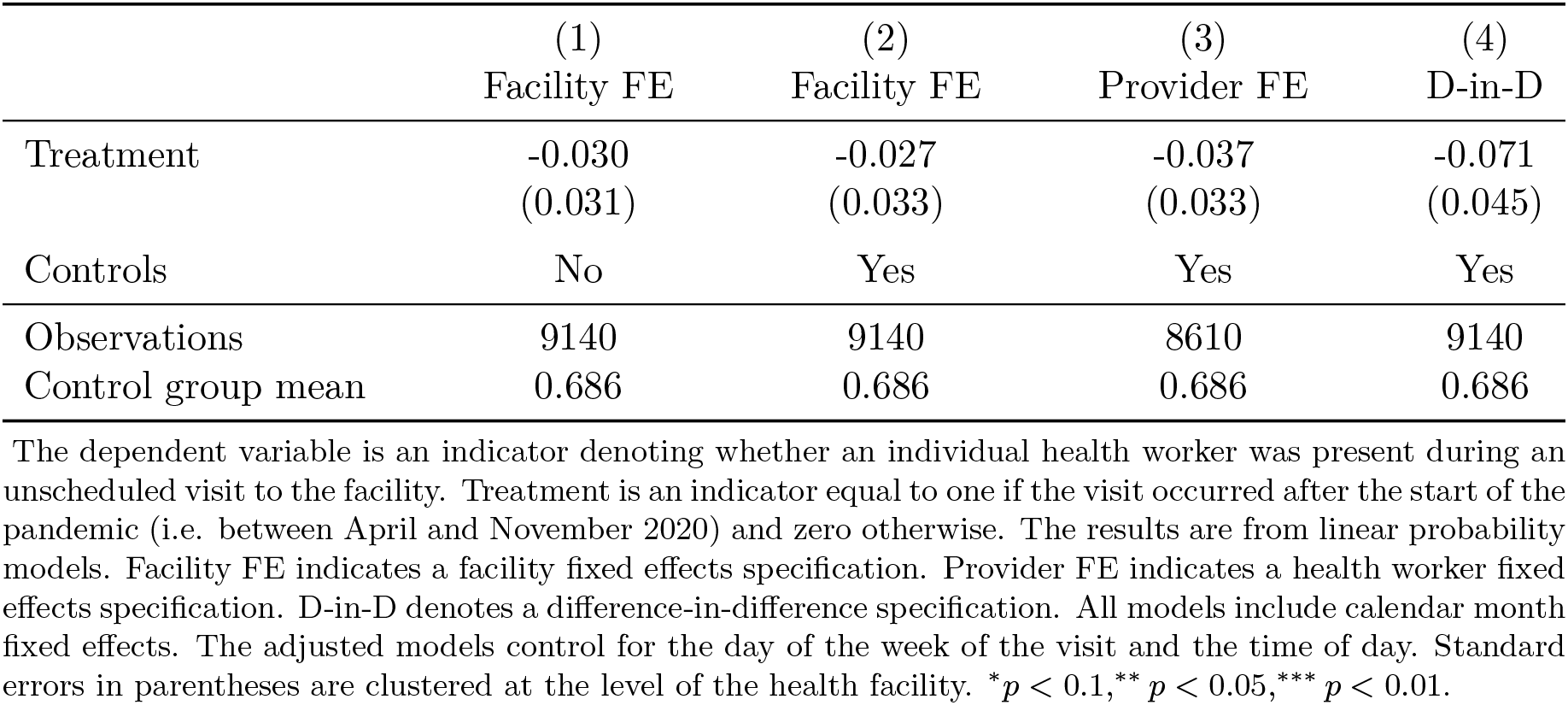
Health Worker Availability

Even though workers continued to come to work did they adjust their behavior along other dimensions? Specifically, I examine whether their interactions with patients changed during the pandemic in ways consistent with risk avoidance. Figure 6a graphs trends in the probability that a sick patient visiting the health facility for care was asked recommended history-taking questions by the health worker. For patients presenting with fever as one of the complaints (68% of patients) there were five recommended questions that we asked about, for patients presenting with a cough (17% of patients) there were five questions, and for patients with diarrhea (4.5% of patients) there were six questions. For all other sick patients, we know if the provider asked the patient whether they were taking any medications or had received any treatment for the complaint. I aggregate all of the responses into a single index by taking an average. The resulting index should be interpreted as adherence to (history-taking) clinical guidelines.

**Figure 6:**
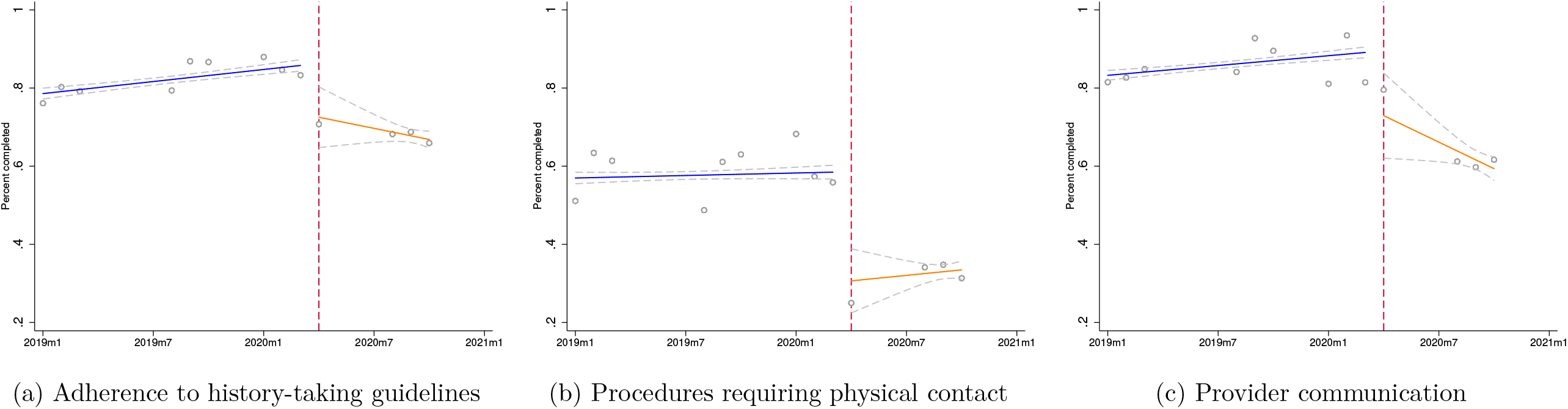
Quality of patient interaction before and during the pandemic Note: Figure 6a graphs the fraction of recommended history-taking questions that were asked by the health worker. Figure 6b graphs the fraction of basic procedures requiring physical contact (a physical examination, a blood pressure check, a temperature check, and an examination with a stethoscope) completed for each patient during the visit. Figure 6c examines the quality of provider communication (the probability that the health worker explained their diagnosis to the patient, provided health education, or discussed when the patient should return for a follow-up). Data are from patient surveys conducted in 288 primary health facilities. Patients were interviewed after the consultation. Circles denote monthly averages. Months with fewer than 25 observations are omitted. The number of observations per month is shown in Table A.1. I fit separate linear regressions to the data before and after the start of the pandemic. Dashed line denotes the start month of the pandemic in Nigeria.

One can see that there was a significant drop in adherence during the pandemic. Adherence, on average, decreased by 16 percentage points; from 84.2% between January-March 2020, to 68.1% between April-November (*P < .*001). In 2019, over the same period, it increased by 6 percentage points (*P* = 0.004). The corresponding regression results in Table 5 Panel A confirm this finding. The estimates of the decrease in adherence range from 17 percentage points in the covariate-adjusted facility fixed effects specification (a relative decrease of 21%), to 21 percentage points in the double-difference specification (a relative decrease of 25%). Results from the health worker fixed effects specification are reported in Appendix Table A.6. All of the results carry through. To clarity for the reader, the sample in Panel A consists of sick patients, in Panel B it includes all patients (sick patients + patients visiting the facility for other kinds of care, e.g., treatment of a minor injury, prenatal care or postnatal care), in Panel C it consists of acute-care patients (sick patient + patients with a minor injury). This is why the sample size varies between panels.

**Table 5:**
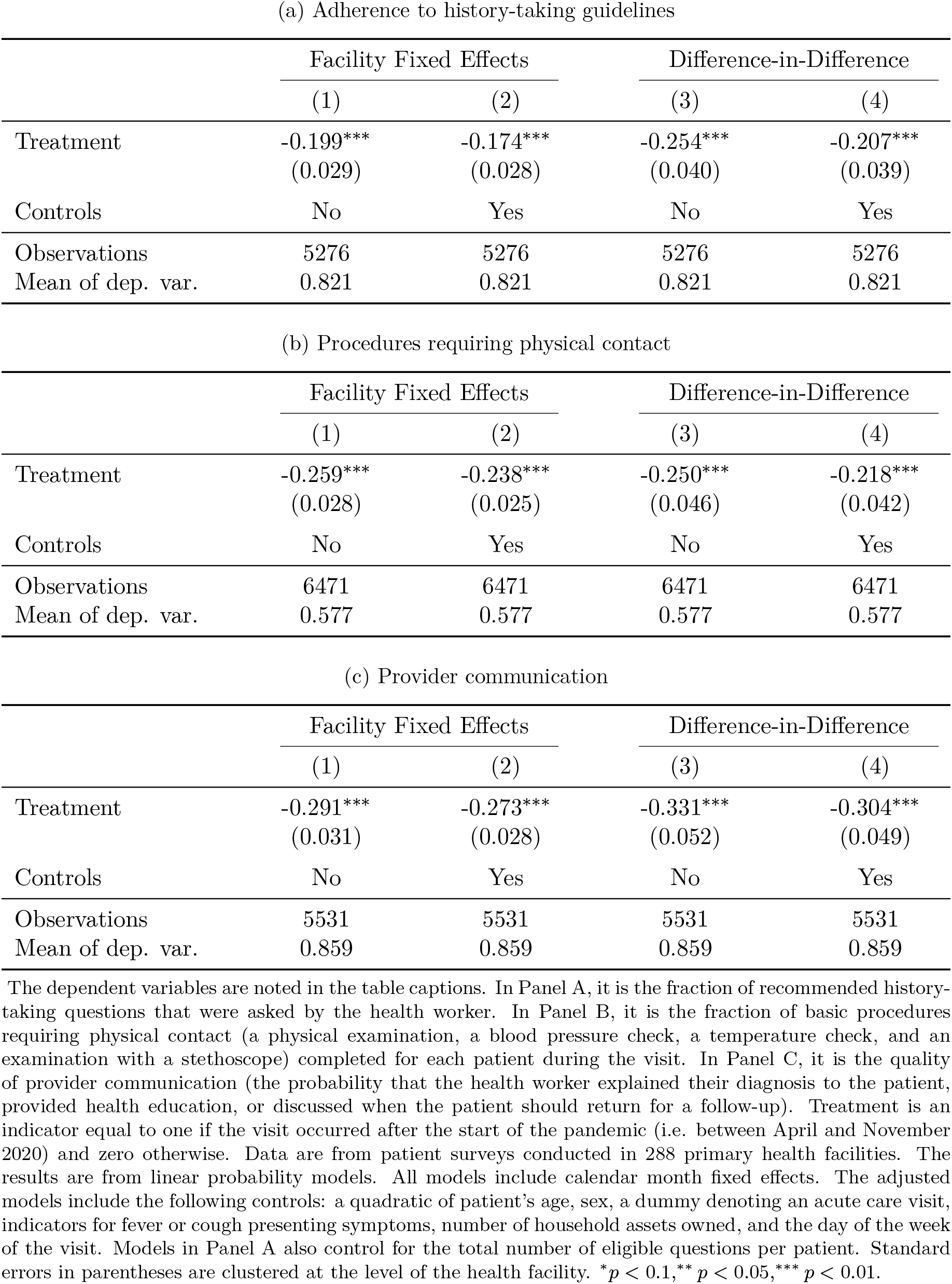
Quality of interaction with patients

Figure 6b examines the probability that a patient received the following procedures that require physical contact with the patient: a physical examination, a blood pressure check, a temperature check, and an examination with a stethoscope. Again, I aggregate by taking an average. A note that I have this information for all patients in the sample: including sick patients and those who visited the health facility for other reasons. 26% of patients did not receive any of these procedures during their visit while 21% received all four (the median is two). We see that the probability that a patient received these procedures fell during the pandemic by 26 percentage points; from 57.3% between January-March 2020, to 33.4% between April-November (*P < .*001). In 2019, over the same period, it increased by 2.2 percentage points (*P* = 0.37). The corresponding regression results in Table 5 Panel B confirm this. The estimates of the decrease range from 22 percentage points in the covariate-adjusted double-difference specification (a relative decrease of 38%), to 24 percentage points in the facility fixed effects specification (a relative decrease of 45%). Results from the health worker fixed effects specification are reported in Table A.6. Again, the results carry through. In Table A.7 I report the results for each individual indicator.

Figure 6c looks at the quality of provider communication. I aggregated the individual indicators into a communication index by taking an average. The results for each individual indicator are in Table A.7. As with other measures of the quality of the clinical interaction, provider communication also reduced significantly. Between January-March and April-November 2020, the probability that the health worker explained their diagnosis to the patient, provided health education, or discussed when to return fell by 25 percentage points (*P < .*001), compared to an increase of 8 percentage points (*P < .*001) over the same period in 2019. The regression results in Table 5 Panel C indicate a decrease of between 27 percentage points in the covariate-adjusted double-difference specification (a relative decrease of 32%), and 30 percentage points in the double-difference specification (a relative decrease of _35%)._18

The results in Table 5 provide persuasive evidence of risk compensation by health workers along the intensive margin. To try to put the results into context, prior to the pandemic nearly 3 in 4 patients received a physical examination (again, this is defined pretty loosely as whether the provider touched the patient) but since the start of the pandemic less than half of patients are receiving an examination. I point out that these changes are not due to pandemic-induced changes in patient composition (controlling for patient characteristics does not alter the results), nor are they due to changes in health worker composition (the results from the health worker specification, despite the differences in the samples, all carry through). A unifying theme for these changes is that they are consistent with health workers attempting to curtail their interactions with patients: asking fewer questions, cutting back on procedures that require physical contact, and providing less health information.

Are there other plausible explanations for these results? I examine this next. One possibility is if patient volume increased during the pandemic. If there were significantly more patients needing care, health workers might hasten their consultations in order to be able to attend to everyone.^19^ I look at this using two different metrics to eliminate all doubt. First, I examine whether the number of patients waiting for service in the waiting area (as recorded by the research assistant during audit visits) noticeably increased during the pandemic. Second, I use monthly patient counts as recorded in facility patient registers. Both variables are log transformed.^20^ There is no evidence of this in Figure A.2. The corresponding regression results are in Table A.8. The results indicate that we can rule out an increase in patient volume as an explanation for these findings. Another possibility is whether these observed changes might be related to the patient interview process itself. If, for example, the research assistants completed these interviews quickly to limit contact, and were perhaps more careless, that could potentially influence the results. We can quickly dismiss this. Using the time stamps on these surveys to measure the duration of the patient interview, Figure A.3 shows that the duration of patient interviews conducted during the pandemic did not decrease.

As a falsification test, I examine whether other dimensions of the interaction, that did not necessarily put the provider at risk by prolonging contact, exhibited a similar decrease. We would not expect them to. In Table A.9 I examine the probability that the provider ordered a lab test and prescribed any medicines. In both cases patients could be asked to wait outside and scripts brought to them. Lab tests are also done by someone else (and somewhere else), and so posed no danger to the health worker; the results could be reviewed by the provider, and any medicines prescribed, without the patient being in the room. As a caveat, I note that nearly all patients were prescribed medicines, so there is a bit of a ceiling effect but I am testing whether there was a decrease. In all cases, I do not find evidence of a decrease along these dimensions.

A final way to test whether these decreases are indicative of risk avoidance is to examine whether they were more likely to affects individuals who, from the perspective of the health worker, posed a greater risk. Patients with symptoms associated with Covid-19 obviously posed a greater risk. Fever and cough were the two symptoms most frequently associated by health workers with Covid-19 (see Figure A.4). 88.7% and 96.4% of health workers surveyed mentioned fever and cough, respectively, as a symptom of Covid-19.^21^ Figure 7 examines the probability that a patient received contact-intensive procedures (a physical exam, blood pressure check, temperature check, and a stethoscope exam) by whether they presented with fever or cough (62% of all patients visiting the health facility presented with at least one of these symptoms). I use the same index in Figure 6b.

**Figure 7:**
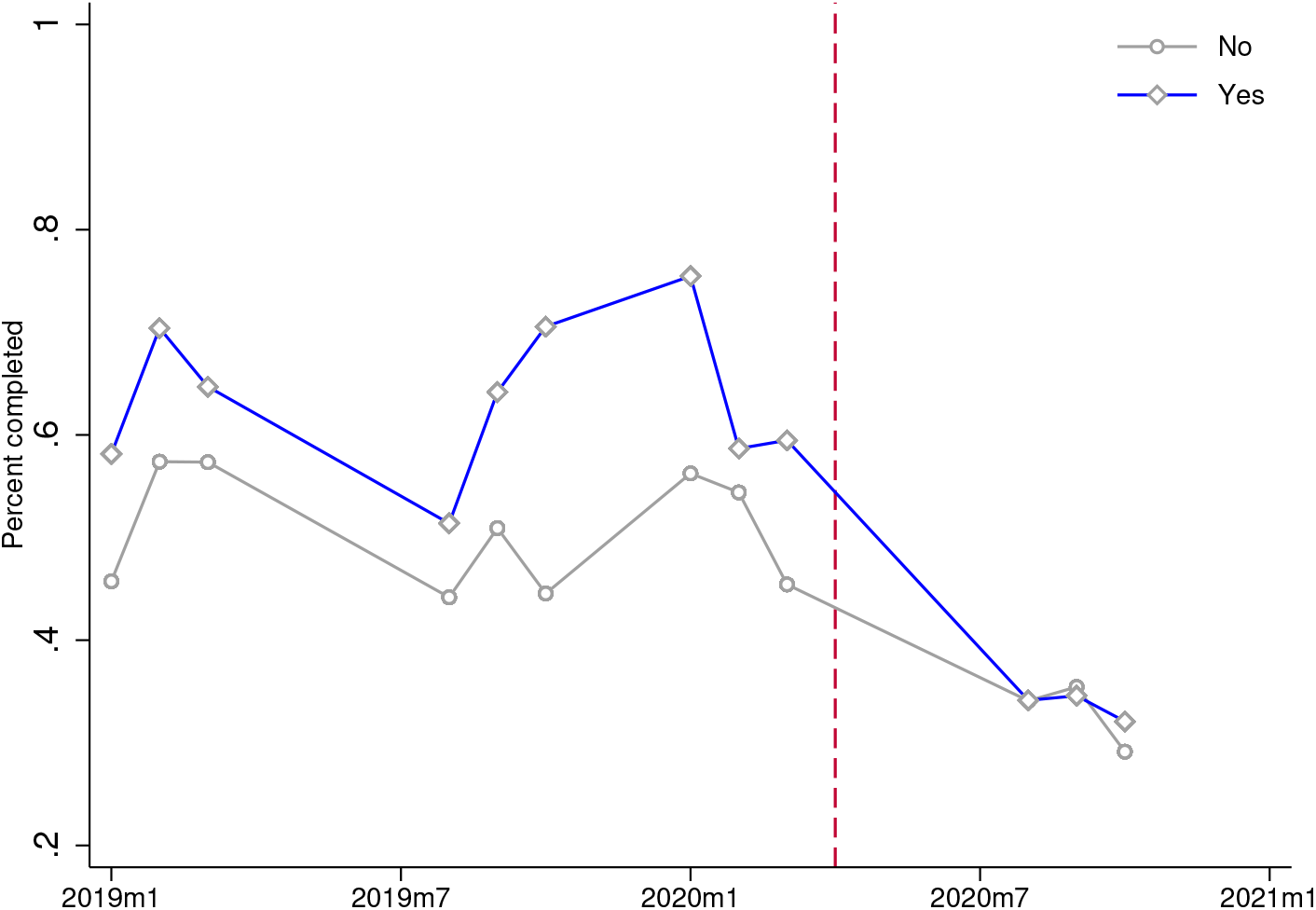
Probability that a patient received procedures requiring physical contact: Patients with/without symptoms associated with Covid-19 Note: Figure graphs the fraction of basic procedures requiring physical contact (a physical examination, a blood pressure check, a temperature check, and an examination with a stethoscope) completed for each patient during the visit. Separate trends are shown for patients presenting with fever or cough, the two symptoms most commonly associated with Covid-19. Data are from patient surveys conducted in 288 primary health facilities. Patients were interviewed after the consultation. Circles denote monthly averages. Months with fewer than 25 observations are omitted. The number of observations per month is shown in Table A.1. Dashed line denotes the start month of the pandemic in Nigeria.

The results are quite striking: while the probability of these procedures fell for all patients, which makes sense since even patients presenting without fever or cough might still have the disease, they fell more sharply for patients with Covid-related symptoms. Prior to the pandemic patients with fever or cough were about 11 percentage points more likely to receive contact-intensive procedures; after the start of the pandemic the rates fell so sharply for patients with fever or cough that they converged with that of patients without these symptoms. In Table 6 I formally test for an interaction effect. I find that, consistent with the visual evidence in Figure 7, these risk avoidance behaviors were most prominent for patients with the symptoms most closely associated with Covid-19. I estimate the same models in Table 5 but interact the treatment indicator with an indicator denoting patients that complained of fever or cough. Across all specifications, the interaction effects are negative and statistically significant, confirming that the incidence of these behaviors fell most heavily on the patients who, from the perspective of the health worker, were the most likely to be infected. However, it is worth stressing that it affected all patients including those without these symptoms (the main effect remains significant). These large negative impacts on patient care raise the obvious question of health implications. I examine this next.

**Table 6:** Quality of patient interaction: Patients with/without symptoms associated with Covid-19

(a) Adherence to history-taking guidelines
|  | Facility Fixed Effects |  | Difference-in-Difference |  |
| --- | --- | --- | --- | --- |
|  | (1) | (2) | (3) | (4) |
| Treatment x COVID | -0.148***<br>(0.044) | -0.091**<br>(0.041) | -0.135***<br>(0.043) | -0.085**<br>(0.041) |
| Treatment | -0.093**<br>(0.043) | -0.109***<br>(0.041) | -0.150***<br>(0.052) | -0.143***<br>(0.051) |
| Controls | No | Yes | No | Yes |
| Observations | 5276 | 5276 | 5276 | 5276 |

(b) Procedures requiring physical contact
|  | Facility Fixed Effects |  | Difference-in-Difference |  |
| --- | --- | --- | --- | --- |
|  | (1) | (2) | (3) | (4) |
| Treatment x COVID | -0.166***<br>(0.033) | -0.154***<br>(0.031) | -0.170***<br>(0.032) | -0.157***<br>(0.030) |
| Treatment | -0.136***<br>(0.032) | -0.122***<br>(0.029) | -0.101**<br>(0.044) | -0.096**<br>(0.041) |
| Controls | No | Yes | No | Yes |
| Observations | 6471 | 6471 | 6471 | 6471 |

(c) Provider communication
|  | Facility Fixed Effects |  | Difference-in-Difference |  |
| --- | --- | --- | --- | --- |
|  | (1) | (2) | (3) | (4) |
| Treatment x COVID | -0.107***<br>(0.038) | -0.094***<br>(0.036) | -0.105***<br>(0.038) | -0.092**<br>(0.036) |
| Treatment | -0.204***<br>(0.034) | -0.198***<br>(0.033) | -0.238***<br>(0.052) | -0.228***<br>(0.051) |
| Controls | No | Yes | No | Yes |
| Observations | 5531 | 5531 | 5531 | 5531 |

### 5.3 Health implications

Figure 8 provides a descriptive look at postpartum complications. This motivates the event study design. The figure shows the eight months before and after the start of the pandemic.^22^ Visually at least, we see evidence of an increase in postpartum complications during the pandemic. Postpartum bleeding increased by 2.2 percentage points; from 15.8% between January-March 2020, to 18% between April-November (*P* = .005). Postpartum infections increased by 5 percentage points; from 6.6% between January-March 2020, to 11.8% between April-November (*P < .*001). The regression results in Table 7 confirm the descriptive results. They indicate that postpartum bleeding and infections increased by 2.5 and 3.7 percentage points, respectively, in the covariate-adjusted models. Including covariates hardly affects the point estimates. Figure A.6 provides empirical validation for the design, showing that maternal characteristics vary smoothly across the cutoff. Importantly, the probability that a woman experienced postpartum bleeding during her last delivery prior to enrollment does not differ.

**Figure 8:**
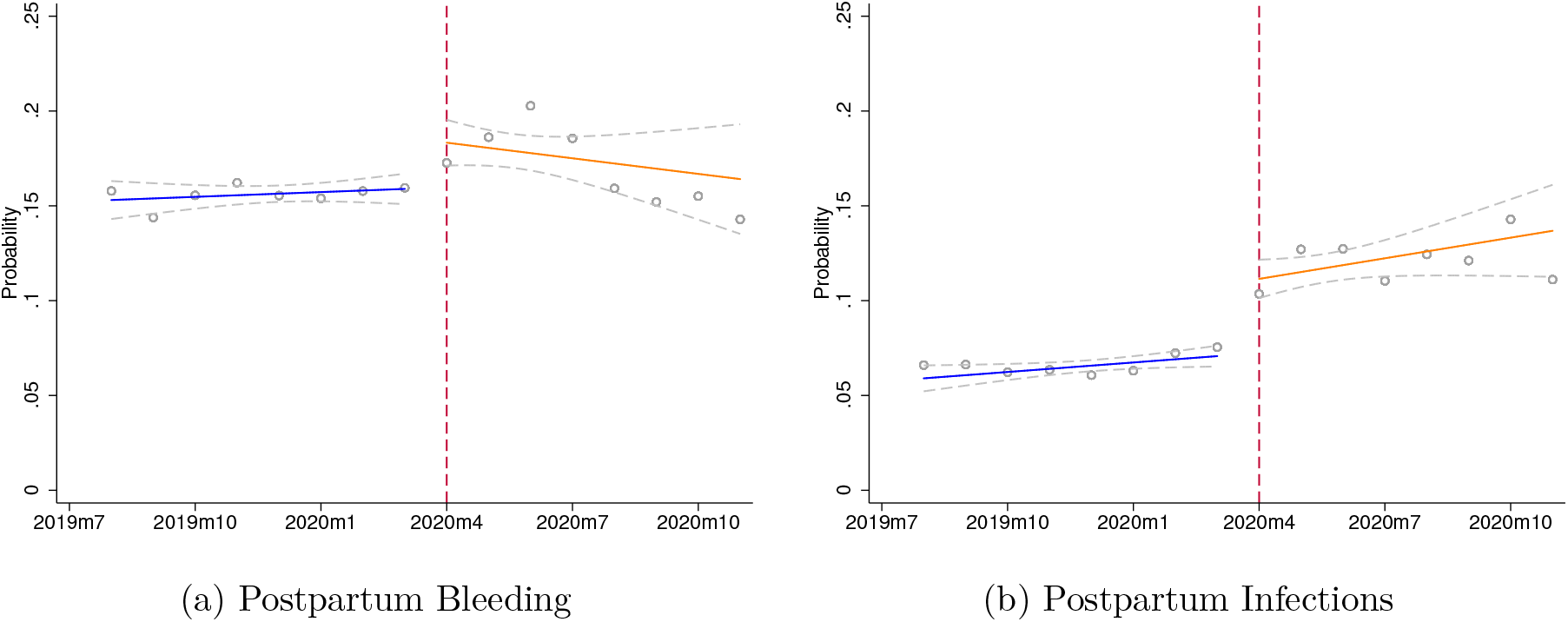
Maternal delivery complications in the 8 months before/after the start of the pandemic Note: Figure graphs the probability of a postpartum complication by month of birth. Data are from an in-home survey of participating women. The women were randomly sampled from communities served by the 288 participating health facilities. The surveys were administered approximately four months after the delivery. The number of deliveries in each month is shown in Table A.10. Circles denote monthly averages. I fit separate linear regressions to the data before and after the start of the pandemic. Dashed line denotes the start month of the pandemic in Nigeria.

**Table 7:**
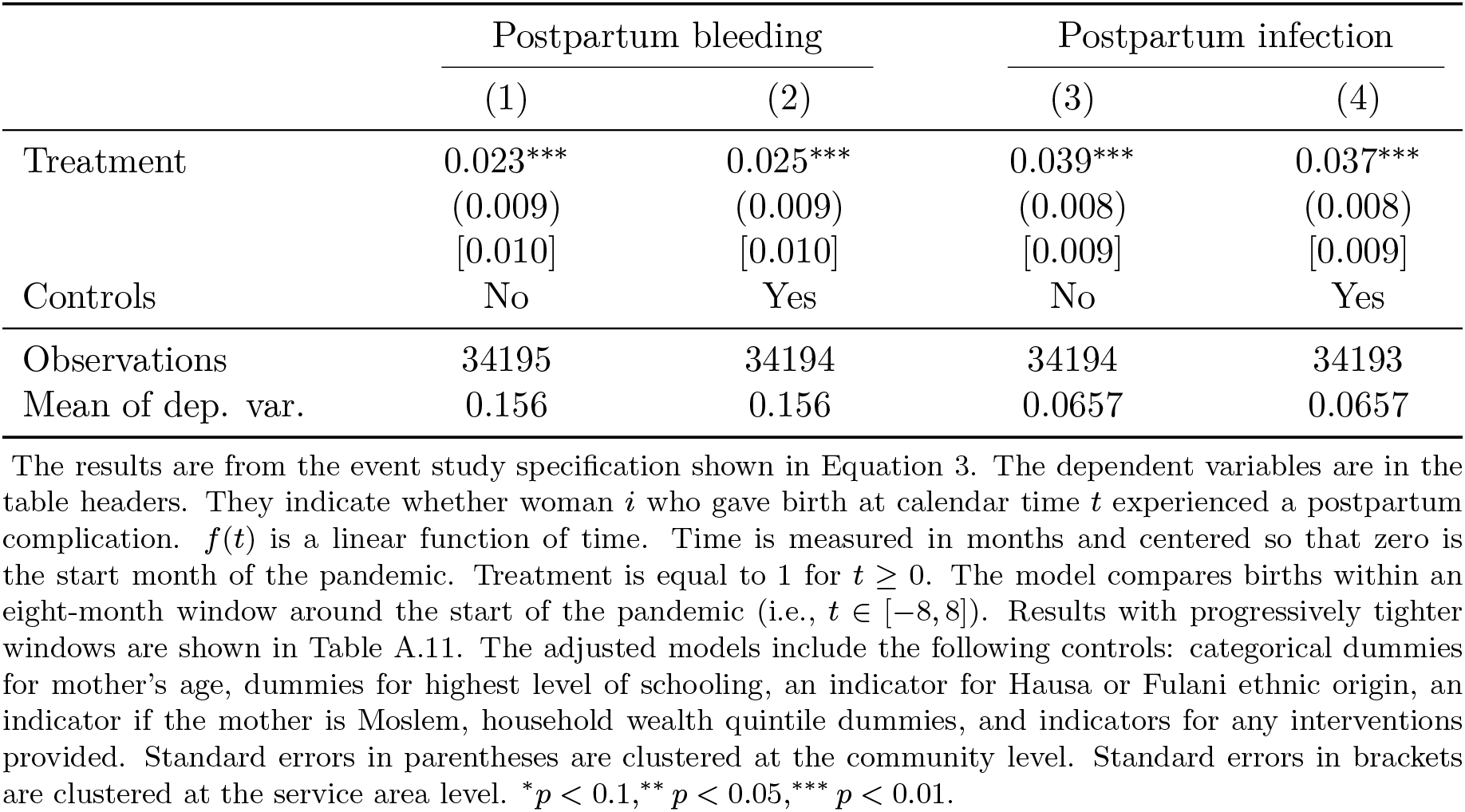
Postpartum Complications: Event Study Models

The results from the robustness checks are reported in the Appendix. Table A.11 examines robustness to tightening the comparison window. I compare deliveries within a six-month, and then a four-month window. The results are stable. Table A.12 shows that the results are largely robust to changes in model specification. Table A.13 presents results from an alternative difference-in-difference model that relies on different identification assumptions. The results are similar to the event study model. Finally, Table A.14 uses the historical data to carry out placebo tests. The basic idea is simple: I re-assign the pandemic to a previous year and re-estimate the same event study specification in Table 7, comparing births within an eight-month window. I iteratively perform the placebo test using April 2017 (Column 1) and April 2018 (Column 2) as the pandemic start date.^23^ Since the pandemic did not occur in those years, we should see no effect on postpartum complications. The results pass these tests as well. These robustness checks indicate that the increase in delivery complications during the pandemic is real and not some quirk of the data. I find no evidence of heterogeneity across intervention groups. The p-values from tests that the interactions terms are jointly equal to zero are 0.4 and 0.8, respectively, for postpartum bleeding and infections.

These results show that postpartum complications increased during the pandemic; whether or not they are solely due to worse care provided during the pandemic is open to some debate. It is certainly possible that there are other mechanisms at work, a point that I will discuss more in the next section. I do not have the right data to definitively establish mechanisms so these results should be taken as suggestive. However if, as the earlier results have indicated, health workers limited their contact with women during labor/delivery, one place where this potentially might show up is in women’s reports about mistreatment during delivery. Women were asked if they were verbally or physically mistreated by health workers during their delivery. About 6% of women reported mistreatment before the pandemic. Table A.15 examines whether reports of mistreatment increased during the pandemic. The results indicate that they did. The incidence increased by 1.4 percentage-points (a relative increase of about 23%). This is a suggestive result that accords with the idea that health workers may have paid less attention to/taken less care of women during delivery.

## 6 Discussion

This study has documented some of the first systematic evidence of risk avoidance behavior by health workers during the Covid-19 pandemic, and how this has impacted patient care and outcomes. Drawing on data collected in 288 frontline primary health facilities in Nigeria over a two-year period from 2019 to 2020, this paper has shown that the quality of health worker interactions with patients has changed in measurable (and predictable) ways during the Covid-19 pandemic. Notably, I found that the probability that patients received contact-intensive procedures such as physical examinations has dramatically declined. The magnitude and breadth of these changes is alarming, especially considering that baseline levels of quality were not very high to begin with. Even more concerning, the data suggest that it could be a while before things to return to normal. It will be crucial to continue to monitor these outcomes as the pandemic continues.

This paper has argued, and presented evidence, that these changes are indicative of risk compensation by health workers along the intensive margin. Since any patient could potentially be carrying the virus, health workers appear to have responded by reducing contact with *all* patients. This kind of risk avoidance behavior may have been exacerbated by the lack of access to Covid testing and lack of high-quality protective equipment. None of the health facilities in the sample were able to conduct Covid-19 testing and only 3% reported having N95 masks in stock (though many health workers supplied their own masks). However, it is important to stress that such behavior is not likely to be limited to only resource-poor settings. Even in high-income countries, many health facilities still struggle to procure enough protective equipment. Additionally, while protective equipment may reduce risk it does not eliminate it, so that health workers are likely to take other measures to protect themselves.^24^

Research in other settings will be useful. Interestingly, I found no evidence of risk compensation on the extensive margin (at least on one dimension, absenteeism). It is possible that this kind of response was limited by moral or ethical considerations (Sahashi et al., 2021) or perhaps by more prosaic considerations such as losing one’s job.

This kind of behavior has obvious health implications. To shed some light on this, this paper examined delivery outcomes for women giving birth during the pandemic and found significantly higher rates of postpartum complications. To be clear, this increase in complications may not be solely due to worse care. There are other possible factors that may have played a role. For example, infection with SARS-CoV-2 may have played a contributory role (Adhikari et al., 2020) or even economic hardships imposed by pandemic-related lockdowns may have impacted women’s health outcomes. However, the principle of Occam’s Razor suggests that the most straightforward explanation is likely to be the correct one. If, as this paper has shown, health workers have responded to the pandemic in ways that restricted access to care and reduced the quality of care being provided, almost certainly, this has contributed to an increase in adverse outcomes. Other possible explanations such as decreased utilization of pregnancy and delivery care during the pandemic are not supported by the data (see Figure A.6).

The findings in this paper run counter to the narrative that poorer countries have largely escaped relatively unscathed from the Covid-19 pandemic (Leonhardt, 2021). While it may be true that some of these countries have not seen the large number of Covid-19-related hospitalizations and deaths seen in Europe and the United States—though there is some evidence that we are probably undercounting deaths (Mwananyanda et al., 2021)—given the already weak health systems that existed in these countries prior to the pandemic, it was inevitable that there would be wider consequences. One of the hallmarks of vulnerability is that even a small shock can precipitate catastrophic consequences. The Covid-19 pandemic is a once-in-a-generation kind of shock. Others have previously warned about secondary or collateral effects of the pandemic (Okeke, 2021; Green, 2020). What this papers shows is that these effects may cut deeper than even pessimistic observers may have suspected. The pandemic remains far from over.

What can we learn from this? In order for health workers to do their jobs adequately, they must feel protected (Imai, 2020). This takes on literal connotations in the context of a life-threatening contagious disease outbreak. While the health care profession may disproportionately attract altruistic individuals (Kolstad and Lindkvist, 2012), altruism has its limits. Health workers are rational individuals and will react in predictable ways to incentives. As we have shown, they will take steps to mitigate their risk when there is a credible threat to their safety. Each individual worker may believe that the potentially negative consequences of their actions are small, but in aggregate these individual actions can add up to quite dramatic effects. This underscores a crucial point: protecting health workers during a pandemic is not only morally justified, it is also smart policy. When thinking about additional spending amidst competing priorities, policymakers must take these effects into account.

The findings in this study call for urgent and immediate policy action. The first priority is to reach out to health providers and, collaboratively, figure out how to improve these outcomes. Punitive strategies are likely to be counter-productive. It is likely that any real solutions will require additional spending, but the cost of doing nothing may be larger still. Even within existing budget constraints, there is likely more than can be done to improve outcomes. For example, policymakers may be able to get some mileage out of giving health providers accurate information about the virus so that they can re-optimize (Akesson et al., 2020; Banerjee et al., 2020). Information interventions are fairly low-cost to provide. Also it goes without saying that, as vaccines become available, health workers should be at the front of the line. At this time it is not clear how willing they will be to take the vaccine, and the likely effects of vaccination on their behavior will depend on how much faith they have in the effectiveness of the vaccine. It does not necessarily follow that if all health workers get vaccinated, that will eliminate all risk avoidance behavior. I leave these issues for future research.

## 7 Conclusion

Approximately one year has passed since the SARS-CoV-2 virus first appeared on the global stage. The fallout has been severe. In particular, it has placed severe strain on health care systems, disrupting the normal provision of health care services. While the direct health effects of the pandemic are visible and obvious, there is growing evidence that its secondary effects could be even more significant, particularly in low and middle-income countries. This study underscores the importance of building resilient health care systems (Hanefeld et al., 2018; Kruk et al., 2017). Even as the rapid development of vaccines promises a return to normalcy, it is important for policy makers to take the hard-won lessons from this pandemic, which have come at great cost in human suffering, and be better prepared to deal with future public health shocks.

## Data Availability

Replication data will be made available with the published manuscript

## Appendix

**Figure A.1:**
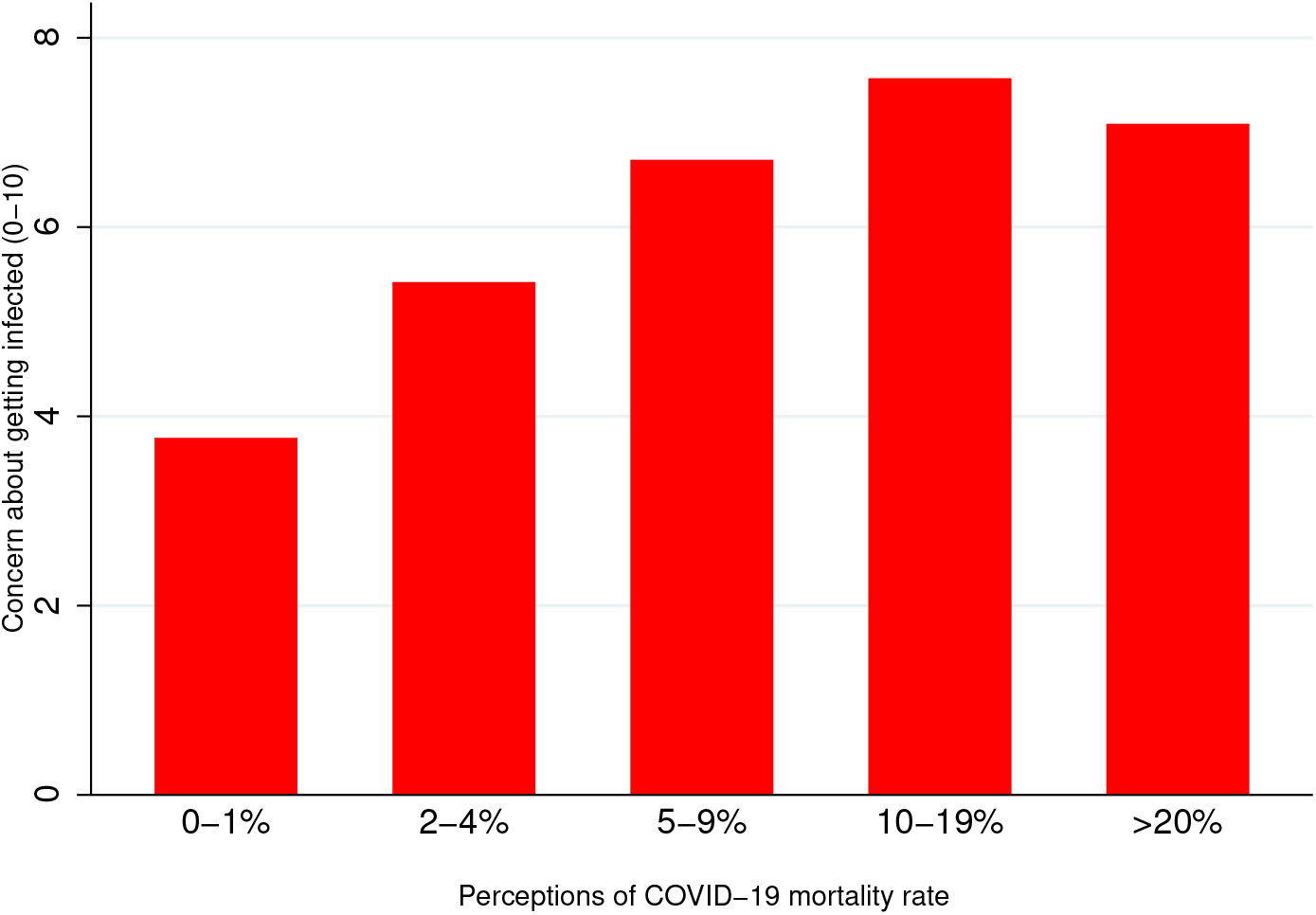
Correlation between health worker beliefs about Covid-19 mortality rate and worry about getting infected. Note: To assess perceptions of Covid-19 mortality, health workers were asked how many people (out of 100) they thought would die if all 100 got infected with the SARS-CoV-2 virus. The same workers were also asked to rate their level of concern about getting infected with Covid-19 on a scale from 0-10, where 0 was no concern at all and 10 was the highest level of concern. Mean scores (on the Y-axis) are computed for various levels of perceived mortality rate (X-axis). The data are from a survey of health workers in 288 primary health facilities in Nigeria that was conducted between August and October 2020.

**Figure A.2:**
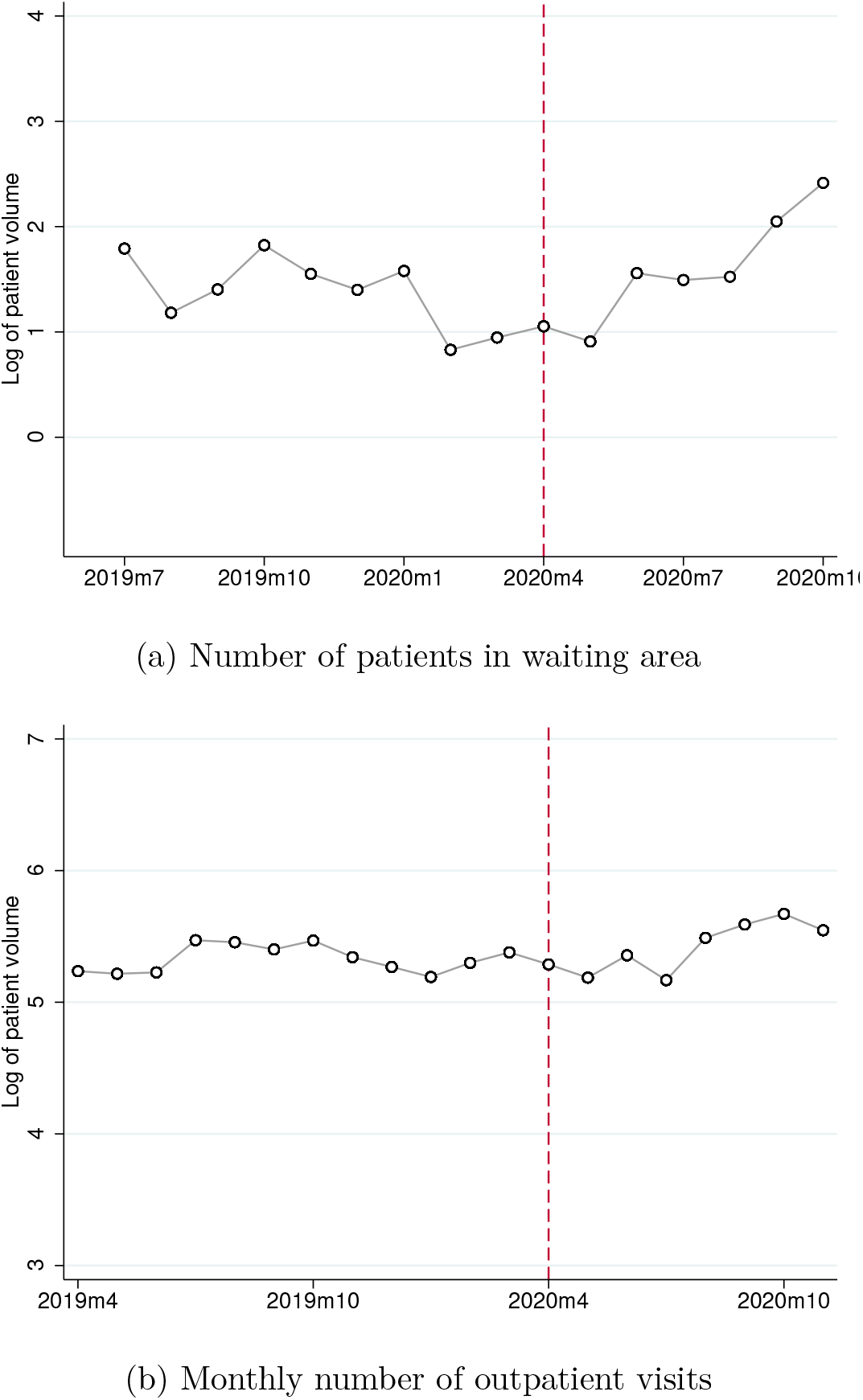
Patient volume before and during the pandemic. Note: Top figure shows trends in the number of patients observed in the waiting area of the health facility. The number of patients in the waiting area was counted during unscheduled visits to the health facility. Monthly averages are shown. The bottom figure shows trends in the monthly number of outpatients seen in the health facility. The data are from health facility patient registers. Both variables are log-transformed. Dashed line denotes the start month of the pandemic in Nigeria.

**Figure A.3:**
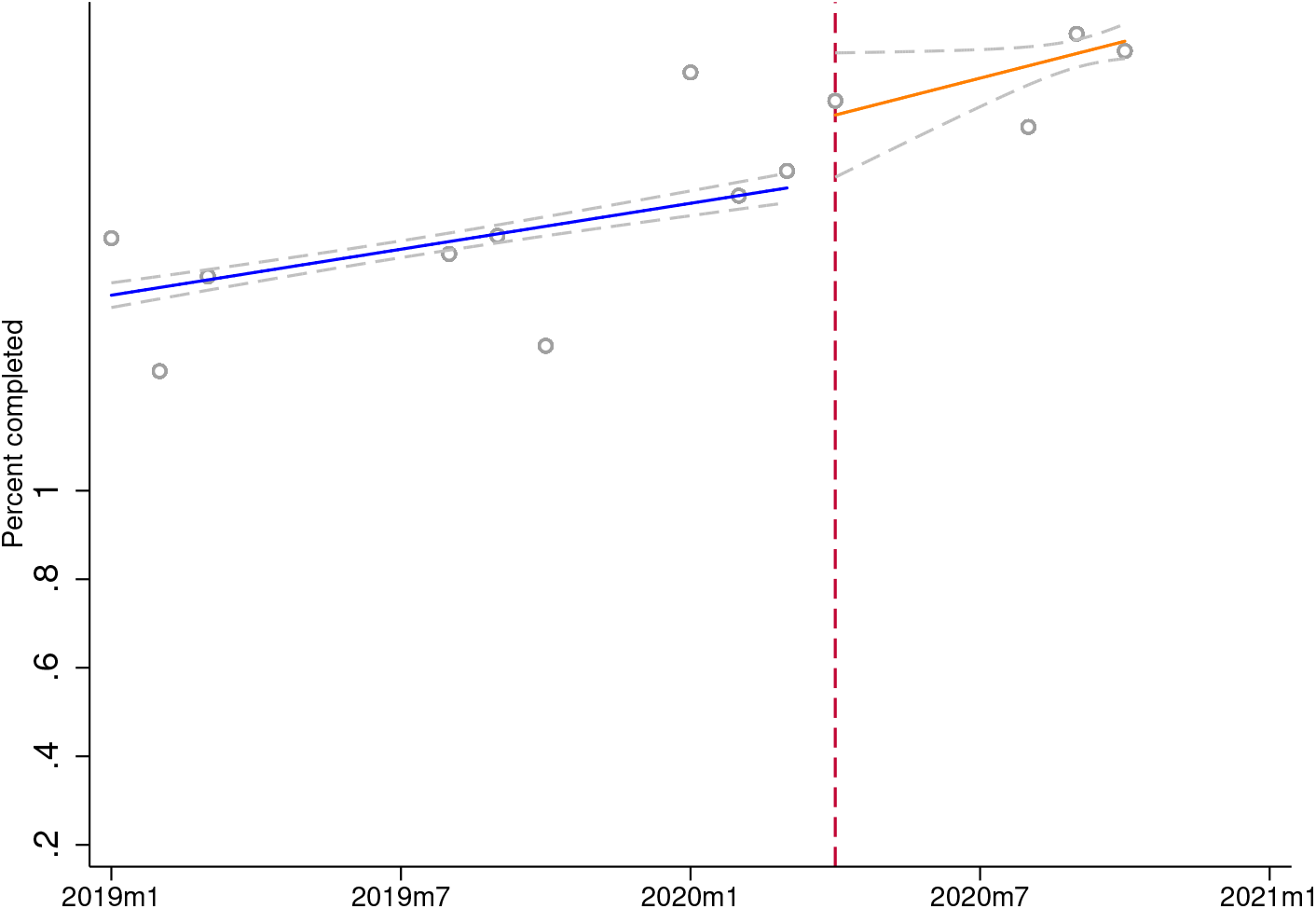
Duration of patient interviews before and during the pandemic. Note: Figure shows trends in the average logged duration of the patient interview. Circles denote monthly averages. Months with fewer than 25 observations are omitted. I fit separate linear regressions to the data before and after the start of the pandemic. Dashed line denotes the start month of the pandemic in Nigeria. Data are from patient surveys conducted in 288 primary health facilities. Patients were interviewed after the consultation. The number of observations per month is shown in Table A.1.

**Figure A.4:**
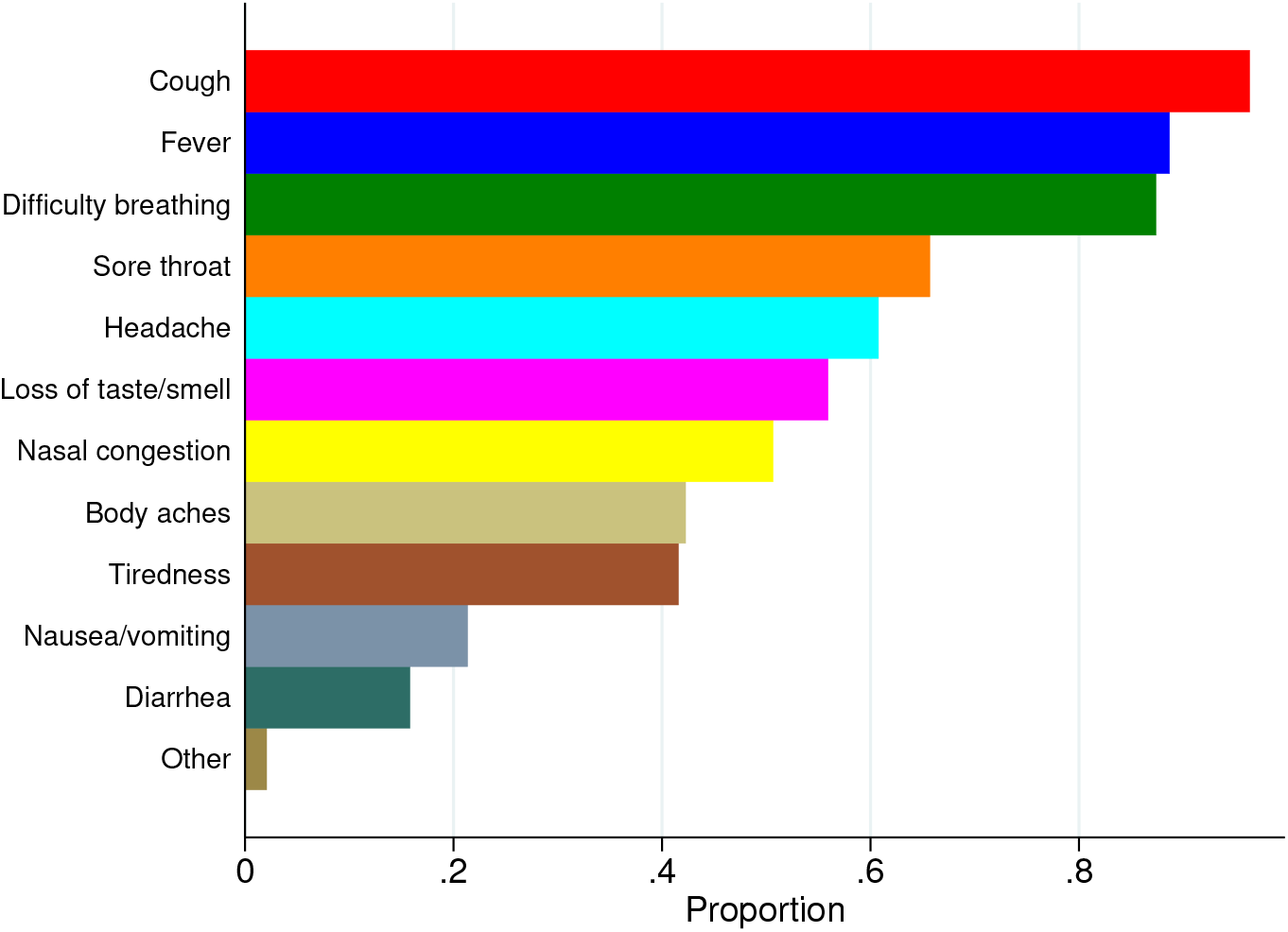
Health worker knowledge of Covid-19 symptoms. Note: Health workers were asked to mention all the symptoms of Covid-19 that they knew. The proportion of health workers mentioning each symptom is shown. The data are from a survey of health workers in 288 primary health facilities in Nigeria that was conducted between August and October 2020.

**Figure A.5:**
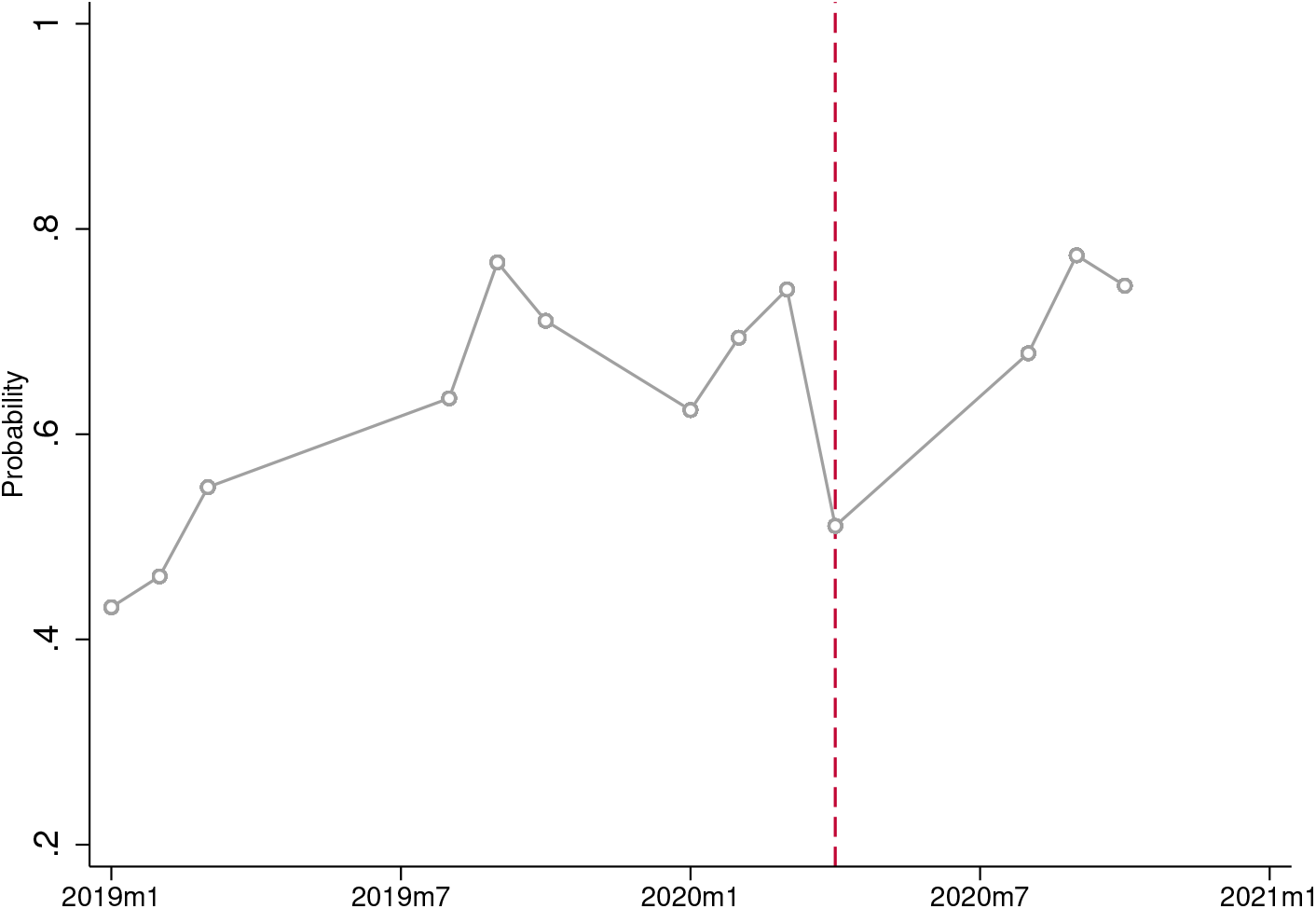
Probability that patient presented with Covid-19 symptoms. Note: Figure shows trends in the proportion of patients presenting with fever or cough, the two symptoms most commonly associated with Covid-19. Dashed line denotes the start month of the pandemic in Nigeria. Data are from patient surveys conducted in 288 primary health facilities. Patients were interviewed after the consultation. The number of observations per month is shown in Table A.1.

**Figure A.6:**
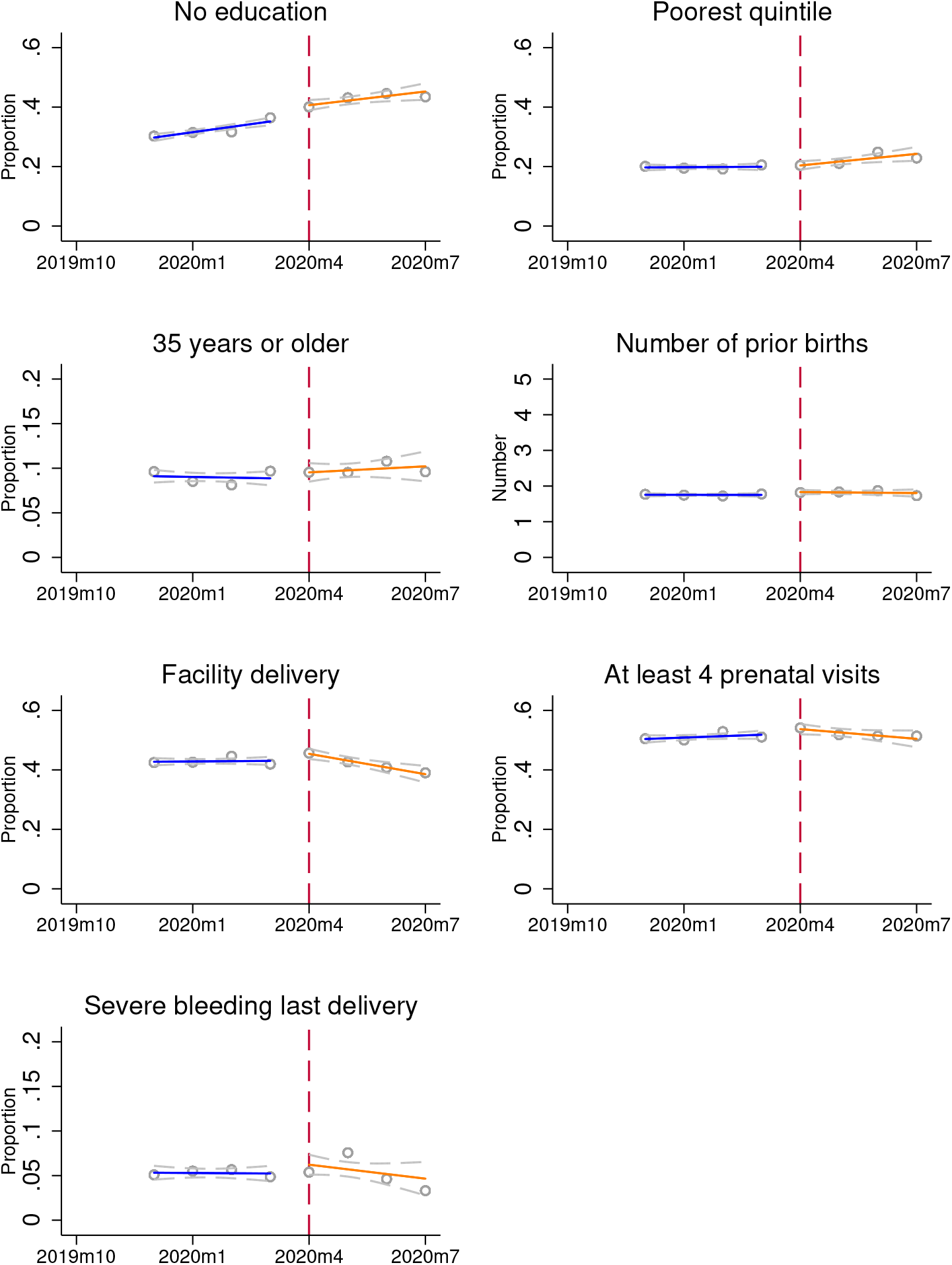
Are there changes in the characteristics of women delivering before and after the start of the pandemic? Note: Data are from an in-home survey of participating women. The women were randomly sampled from communities served by the 288 participating health facilities. Women’s characteristics are from the baseline survey. Utilization variables are from the endline survey administered approximately four months after the delivery. The number of deliveries in each month is shown in Table A.10. Circles denote monthly averages. I fit separate linear regressions to the data before and after the start of the pandemic. Dashed line denotes the start month of the pandemic in Nigeria.

**Table A.1:** Number of patients interviewed each month

|  | Visit month |
| --- | --- |
| Jan 2019 | 1196 |
| Feb 2019 | 791 |
| Mar 2019 | 527 |
| Apr 2019 | 10 |
| Aug 2019 | 200 |
| Sep 2019 | 701 |
| Oct 2019 | 190 |
| Nov 2019 | 16 |
| Jan 2020 | 85 |
| Feb 2020 | 706 |
| Mar 2020 | 676 |
| Apr 2020 | 47 |
| May 2020 | 4 |
| Jul 2020 | 8 |
| Aug 2020 | 137 |
| Sep 2020 | 647 |
| Oct 2020 | 521 |
| Nov 2020 | 9 |
| Observations | 6471 |

**Table A.2:** Number of health worker observations per month

|  | Visit month |
| --- | --- |
| Jan 2019 | 782 |
| Feb 2019 | 466 |
| Mar 2019 | 390 |
| Jun 2019 | 1341 |
| Jul 2019 | 208 |
| Aug 2019 | 425 |
| Sep 2019 | 399 |
| Oct 2019 | 427 |
| Nov 2019 | 203 |
| Dec 2019 | 350 |
| Jan 2020 | 399 |
| Feb 2020 | 604 |
| Mar 2020 | 910 |
| Apr 2020 | 603 |
| May 2020 | 525 |
| Jun 2020 | 635 |
| Jul 2020 | 139 |
| Aug 2020 | 47 |
| Sep 2020 | 198 |
| Oct 2020 | 89 |
| Observations | 9140 |

**Table A.3:** Interrater agreement between patient reports and independent observer reports

| Measure | Agreement | Kappa |
| --- | --- | --- |
| Fever: Asked about pattern | 0.941 | 0.663 |
| Fever: Asked whether vomit | 0.912 | 0.565 |
| Fever: Asked whether chills/sweats | 0.925 | 0.710 |
| Fever: Asked whether catarrh/runny nose | 0.913 | 0.698 |
| Fever: Asked whether convulsions | 0.897 | 0.737 |
| Examined patient | 0.837 | 0.642 |
| Examined patient with a stethoscope | 0.874 | 0.733 |

For each listed variable, the table reports the percent agreement between the patient reports and the report of the independent observer for the same interaction, and the computed kappa statistic. Values between 0.41 and 0.6 are considered moderate agreement, values between 0.61-0.8 are considered substantial agreement, and values between 0.81 and 1 are considered perfect agreement. I focus on fever because this accounts for the vast majority of complaints. Blood pressure and temperature measurement are not used because they often happen outside of the consultation room and thus would not be observed by the independent observer.

**Table A.4:** Characteristics of health workers that completed the Covid-19 survey module

|  | mean | sd |
| --- | --- | --- |
| Age | 37.4 | 8.92 |
| Male | .555 | .497 |
| College-educated | .0582 | .234 |
| Total years of clinical work experience | 12.3 | 7.95 |
| Facility-specific experience | 5.68 | 5.48 |
| Observations | 1324 |  |

The table describes the characteristics of health workers surveyed during Visit 4 (August-October 2020) when the Covid-19 survey module was added to the survey instrument. Facility-specific experience indicates the number of years since they started working in the health facility.

**Table A.5:** Do health worker characteristics predict beliefs?

|  | (1)<br>Perceived Covid<br>mortality | (2)<br>Perceived Covid<br>mortality | (3)<br>Worried about<br>getting Covid-19 | (4)<br>Beliefs about<br>transmission |
| --- | --- | --- | --- | --- |
| Male | 1.765*<br>(0.966) | 0.315***<br>(0.082) | 1.417***<br>(0.210) | -0.905***<br>(0.145) |
| Age | 0.015<br>(0.095) | 0.001<br>(0.008) | -0.032*<br>(0.018) | 0.024*<br>(0.013) |
| College-educated | -1.599<br>(1.855) | -0.279*<br>(0.157) | -0.882**<br>(0.369) | 0.184<br>(0.268) |
| Total years of clinical work experience | -0.218**<br>(0.109) | -0.014<br>(0.009) | 0.002<br>(0.022) | 0.009<br>(0.017) |
| Observations | 1198 | 1198 | 1198 | 1198 |
| Mean of dep. var. | 9.557 | 2.830 | 6.073 | 5.886 |

The dependent variables are in the table header. To assess perceptions of mortality, health workers were asked how many people (out of 100) they thought would die if all 100 got infected with the SARS-CoV-2 virus. In Column 1 the dependent variable is the reported number. Because there is heaping at intervals of 5 (10) below (above) 10 so, in Column 2, the variable is discretized as follows: 0-1, 2-4, 5-9, 10-19, $\geq 20$ . In Column 3 the dependent variable is the level of concern about getting infected. Workers were asked how worried they were about getting infected with Covid-19 on a scale from 0-10, where 0 was no concern at all and 10 was the highest level of concern. In Column 4 the dependent variable is accuracy of beliefs about transmission. I assign a score of +1 for correct responses, -1 for incorrect responses, and 0 for neutral responses and sum. If all responses are correct/incorrect the maximum score is 8/-8. Data are from health worker surveys conducted in 288 primary health facilities. 1,324 workers responded to the survey but 126 reported not being aware of Covid-19 and were not asked these questions. Reported estimates are average marginal effects from Poisson models. Standard errors in parentheses are clustered at the level of the health facility. \* $p < 0.1$ , \*\* $p < 0.05$ , \*\*\* $p < 0.01$

**Table A.6:** Quality of interaction with patients: Health Worker Fixed Effects Specification

|  | History-taking |  | Basic procedures |  | Provider communication |  |
| --- | --- | --- | --- | --- | --- | --- |
|  | (1) | (2) | (3) | (4) | (5) | (6) |
| Treatment | -0.077**<br>(0.038) | -0.074*<br>(0.038) | -0.102***<br>(0.038) | -0.101***<br>(0.034) | -0.132***<br>(0.038) | -0.133***<br>(0.036) |
| Controls | No | Yes | No | Yes | No | Yes |
| Observations | 3337 | 3337 | 3744 | 3744 | 3411 | 3411 |
| Mean of dep. var. | 0.848 | 0.848 | 0.582 | 0.582 | 0.889 | 0.889 |

The dependent variables are in the table headers. History-taking indicates the fraction of recommended history-taking questions that were asked by the health worker. Basic procedures indicates whether the following were completed for each patient during the visit: a physical examination, a blood pressure check, a temperature check, and an examination with a stethoscope. Provider communication indicates whether the health worker explained their diagnosis to the patient, provided health education, or discussed when the patient should return for a follow-up. Treatment is an indicator equal to one if the visit occurred after the start of the pandemic (i.e. between April and November 2020) and zero otherwise. Data are from patient surveys conducted in 288 primary health facilities. The adjusted models include the following controls: a quadratic of patient's age, sex, a dummy denoting an acute care visit, indicators for fever or cough presenting symptoms, number of household assets owned, and the day of the week of the visit. Column 1 also controls for the total number of eligible questions per patient. Standard errors in parentheses are clustered at the level of the health facility. \* $p < 0.1$ , \*\* $p < 0.05$ , \*\*\* $p < 0.01$ .

**Table A.7:** Quality of interaction with patients: Individual indicators

|  | Basic procedures |  |  |  | Provider communication |  |  |
| --- | --- | --- | --- | --- | --- | --- | --- |
|  | (1) | (2) | (3) | (4) | (5) | (6) | (7) |
|  | BP check | Temp check | Physical exam | Used stethoscope | Told diagnosis | Health education | Return visit |
| during | -0.277***<br>(0.040) | -0.156***<br>(0.056) | -0.236***<br>(0.049) | -0.204***<br>(0.056) | -0.362***<br>(0.049) | -0.322***<br>(0.052) | -0.228***<br>(0.056) |
| Controls | Yes | Yes | Yes | Yes | Yes | Yes | Yes |
| Observations | 6471 | 6471 | 6471 | 6471 | 5531 | 5531 | 5531 |
| Mean of dep. var. | 0.501 | 0.565 | 0.689 | 0.552 | 0.881 | 0.877 | 0.821 |

The dependent variables are in the table headers. Basic procedures indicates whether the following were completed for each patient during the visit: a physical examination, a blood pressure check, a temperature check, and an examination with a stethoscope. Provider communication indicates whether the health worker explained their diagnosis to the patient, provided health education, or discussed when the patient should return for a follow-up. Treatment is an indicator equal to one if the visit occurred after the start of the pandemic (i.e. between April and November 2020) and zero otherwise. Data are from patient surveys conducted in 288 primary health facilities. The results are from the difference-in-difference specification shown in Equation 2. All models include the following controls: a quadratic of patient's age, sex, a dummy denoting an acute care visit, indicators for fever or cough presenting symptoms, number of household assets owned, and the day of the week of the visit. Standard errors in parentheses are clustered at the level of the health facility.
\* $p < 0.1$ , \*\* $p < 0.05$ , \*\*\* $p < 0.01$ .

**Table A.8:**
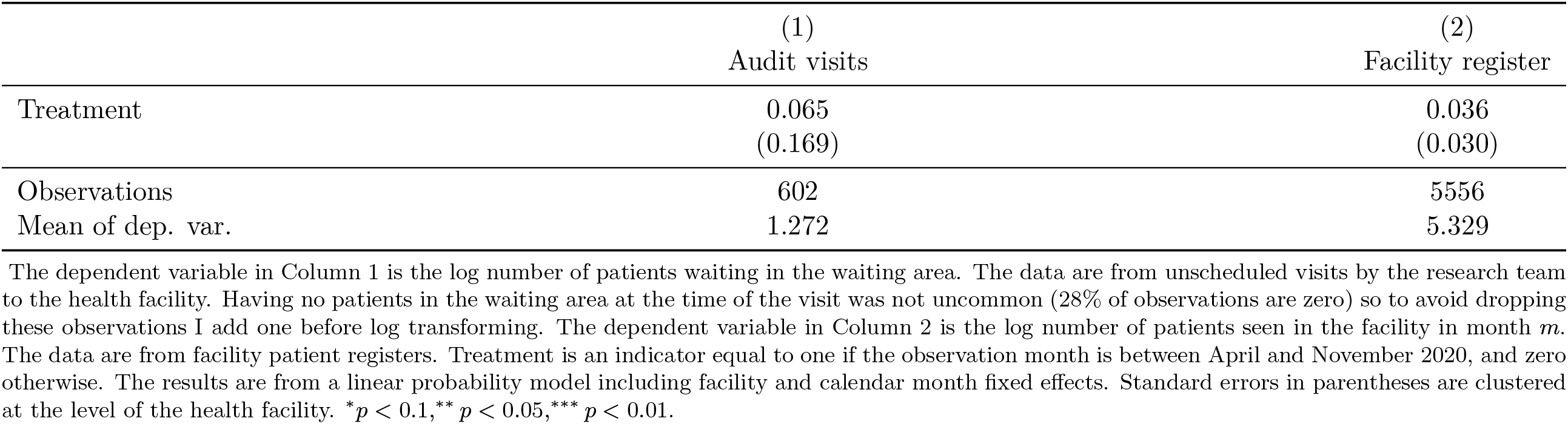
Did patient volume increase during the pandemic?

**Table A.9:** Falsification Tests

|  | Ordered a lab test |  |  | Prescribed medicines |  |  |
| --- | --- | --- | --- | --- | --- | --- |
|  | (1)<br>Facility FE | (2)<br>Provider FE | (3)<br>D-in-D | (4)<br>Facility FE | (5)<br>Provider FE | (6)<br>D-in-D |
| Treatment | 0.045*<br>(0.027) | -0.024<br>(0.043) | -0.034<br>(0.047) | 0.002<br>(0.005) | -0.002<br>(0.006) | -0.001<br>(0.009) |
| Controls | Yes | Yes | Yes | Yes | Yes | Yes |
| Observations | 5531 | 3466 | 5531 | 5531 | 3466 | 5531 |
| Mean of dep. var. | 0.687 | 0.773 | 0.687 | 0.982 | 0.988 | 0.982 |

The dependent variables are noted in the table headers. Lab test indicates whether the health worker ordered a lab test. Medicines indicates whether the health worker prescribed medicines for the patient. Treatment is an indicator equal to one if the visit occurred after the start of the pandemic (i.e. between April and November 2020) and zero otherwise. Data are from patient surveys conducted in 288 primary health facilities. The results are from linear probability models. All models include calendar month fixed effects. All models include the following controls: a quadratic of patient's age, sex, a dummy denoting an acute care visit, indicators for fever or cough presenting symptoms, number of household assets owned, and the day of the week of the visit. Standard errors in parentheses are clustered at the level of the health facility. \* $p < 0.1$ , \*\* $p < 0.05$ , \*\*\* $p < 0.01$ .

**Table A.10:** Number of deliveries per month

|  | Month of birth |
| --- | --- |
| Aug 2019 | 773 |
| Sep 2019 | 2051 |
| Oct 2019 | 3742 |
| Nov 2019 | 4741 |
| Dec 2019 | 4613 |
| Jan 2020 | 4203 |
| Feb 2020 | 3625 |
| Mar 2020 | 3366 |
| Apr 2020 | 2473 |
| May 2020 | 1826 |
| Jun 2020 | 1021 |
| Jul 2020 | 652 |
| Aug 2020 | 402 |
| Sep 2020 | 388 |
| Oct 2020 | 245 |
| Nov 2020 | 63 |
| Dec 2020 | 11 |
| Observations | 34195 |

**Table A.11:** Postpartum Complications: Robustness to shrinking the time window for comparison

|  | Postpartum bleeding |  |  | Postpartum infection |  |  |
| --- | --- | --- | --- | --- | --- | --- |
|  | (1)<br>8 months | (2)<br>6 months | (3)<br>4 months | (4)<br>8 months | (5)<br>6 months | (6)<br>4 months |
| Treatment | 0.025***<br>(0.009) | 0.027***<br>(0.009) | 0.020*<br>(0.010) | 0.037***<br>(0.008) | 0.034***<br>(0.008) | 0.027***<br>(0.008) |
| Controls | Yes | Yes | Yes | Yes | Yes | Yes |
| Observations | 34194 | 31296 | 22180 | 34193 | 31295 | 22180 |
| Mean of dep. var. | 0.156 | 0.157 | 0.156 | 0.066 | 0.066 | 0.067 |

The results are from the event study specification shown in Equation 3. The dependent variables are in the table headers. They indicate whether woman $i$ who gave birth at calendar time $t$ experienced a postpartum complication. $f(t)$ is a linear function of time. Time is measured in months and centered so that zero is the start month of the pandemic. Treatment is equal to 1 for $t \geq 0$ . The model compares births within an eight-month window around the start of the pandemic, then six, and then four months. All models include the following controls: categorical dummies for mother's age, dummies for highest level of schooling, an indicator for Hausa or Fulani ethnic origin, an indicator if the mother is Moslem, household wealth quintile dummies, and indicators for any interventions provided. Standard errors in parentheses are clustered at the community level. \* $p < 0.1$ , \*\* $p < 0.05$ , \*\*\* $p < 0.01$ .

**Table A.12:** Postpartum Complications: Alternative event study specifications

|  | Postpartum bleeding |  | Postpartum infection |  |
| --- | --- | --- | --- | --- |
|  | (1) | (2) | (3) | (4) |
| Treatment | 0.016*<br>(0.009) | 0.020*<br>(0.012) | 0.034***<br>(0.008) | 0.023***<br>(0.009) |
| Controls | Yes | Yes | Yes | Yes |
| Facility FE | Yes | No | Yes | No |
| Trends | Linear | Quadratic | Linear | Quadratic |
| Observations | 34194 | 34194 | 34193 | 34193 |
| Mean of dep. var. | 0.156 | 0.156 | 0.066 | 0.066 |

The results are from alternative specifications of the event study model shown in Equation 3. The dependent variables are in the table headers. They indicate whether woman $i$ who gave birth at calendar time $t$ experienced a postpartum complication. Columns 1 and 3 include service area fixed effects and, in Columns 2 and 4, time is specified as a quadratic. Time is measured in months and centered so that zero is the start month of the pandemic. Treatment is equal to 1 for $t \geq 0$ . The model compares births within an eight-month window around the start of the pandemic. All models include the following controls: categorical dummies for mother's age, dummies for highest level of schooling, an indicator for Hausa or Fulani ethnic origin, an indicator if the mother is Moslem, household wealth quintile dummies, and indicators for any interventions provided. Standard errors in parentheses are clustered at the community level. \* $p < 0.1$ , \*\* $p < 0.05$ , \*\*\* $p < 0.01$ .

**Table A.13:** Postpartum Complications: Difference-in-Difference Estimates

|  | (1)<br>Bleeding | (2)<br>Bleeding |
| --- | --- | --- |
| Treatment | 0.016**<br>(0.008) | 0.019**<br>(0.008) |
| Controls | No | Yes |
| Observations | 52960 | 52955 |
| Mean of dep. var. | 0.115 | 0.115 |

The results are from a difference-in-difference specification that uses retrospective data from women's birth histories. I have data on postpartum bleeding (the dependent variable) for the most recent delivery in the three years preceding enrollment. Treatment is an indicator equal to one if the delivery occurred in or after April in a calendar year, and zero otherwise. All models include community, calendar year, and calendar month of birth fixed effects. The adjusted models include the following controls: categorical dummies for mother's age, dummies for highest level of schooling, an indicator for Hausa or Fulani ethnic origin, an indicator if the mother is Moslem, household wealth quintile dummies, and indicators for any interventions provided. Standard errors in parentheses are clustered at the community level. \* $p < 0.1$ , \*\* $p < 0.05$ , \*\*\* $p < 0.01$ .

**Table A.14:** Placebo tests for postpartum bleeding

|  | (1)<br>2016-2017 | (2)<br>2017-2018 |
| --- | --- | --- |
| Treatment | 0.006<br>(0.007) | -0.010<br>(0.014) |
| Observations | 11976 | 5297 |

The table presents results from placebo tests using data from women's birth histories. I have data on postpartum bleeding (the dependent variable) for the most recent delivery in the three years preceding enrollment (since January 2016). I re-estimate the event study model shown in Equation 3 comparing births within an eight-month window around April 2017 (Column 1) and April 2018 (Column 2). This reassigns the pandemic to 2017 and 2018 respectively. I cannot perform the same test for 2019 because there were only a few births over the relevant time period. In the four months around April 2019 there were only 334 deliveries. All models include the following controls: categorical dummies for mother's age, dummies for highest level of schooling, an indicator for Hausa or Fulani ethnic origin, an indicator if the mother is Moslem, household wealth quintile dummies, and indicators for any interventions provided. Standard errors in parentheses are clustered at the community level. \* $p < 0.1$ , \*\* $p < 0.05$ , \*\*\* $p < 0.01$ .

**Table A.15:**
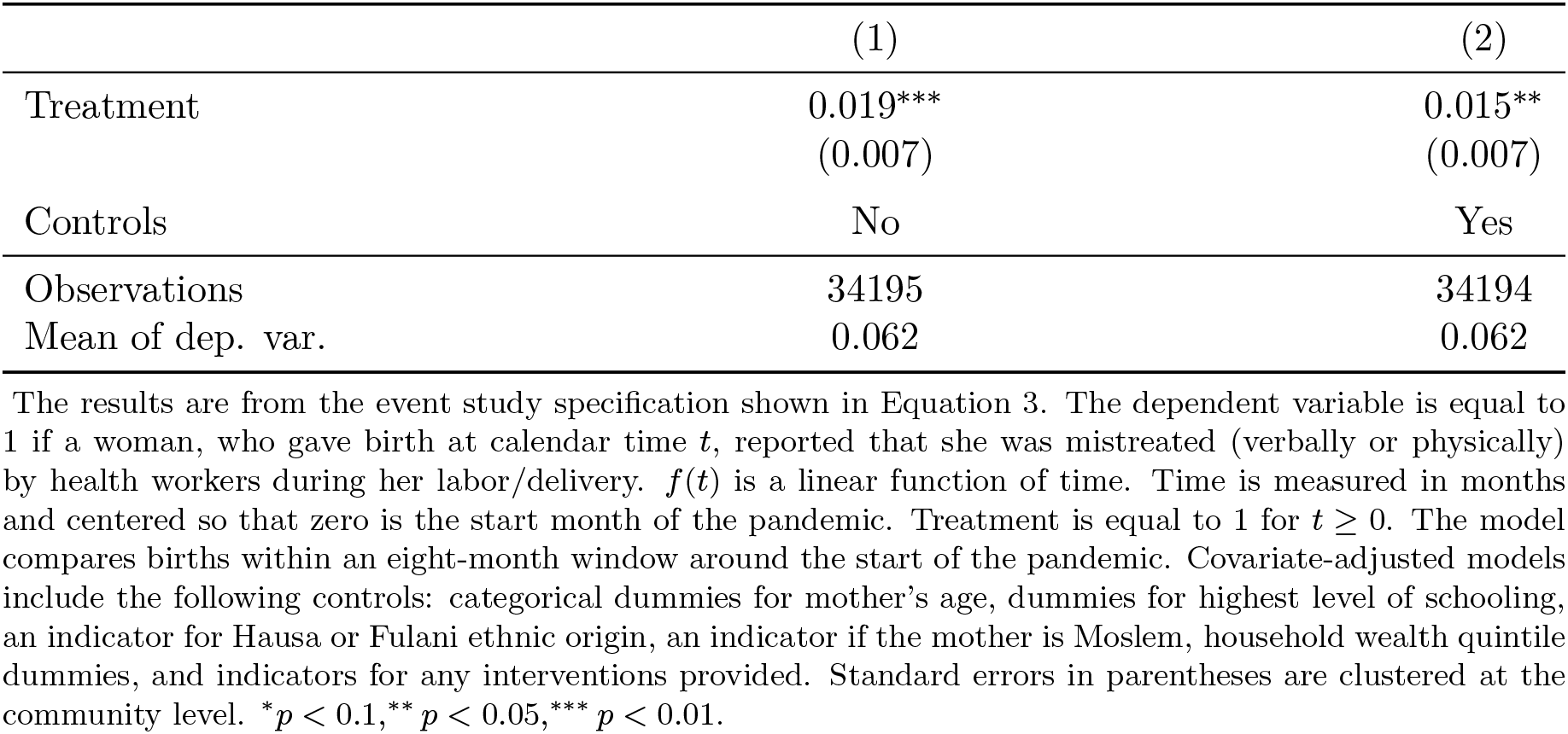
Women reports of mistreatment by health workers during the pandemic

|  | (1) | (2) |
| --- | --- | --- |
| Treatment | 0.019***<br>(0.007) | 0.015**<br>(0.007) |
| Controls | No | Yes |
| Observations | 34195 | 34194 |
| Mean of dep. var. | 0.062 | 0.062 |

The results are from the event study specification shown in Equation 3. The dependent variable is equal to 1 if a woman, who gave birth at calendar time $t$ , reported that she was mistreated (verbally or physically) by health workers during her labor/delivery. $f(t)$ is a linear function of time. Time is measured in months and centered so that zero is the start month of the pandemic. Treatment is equal to 1 for $t \geq 0$ . The model compares births within an eight-month window around the start of the pandemic. Covariate-adjusted models include the following controls: categorical dummies for mother's age, dummies for highest level of schooling, an indicator for Hausa or Fulani ethnic origin, an indicator if the mother is Moslem, household wealth quintile dummies, and indicators for any interventions provided. Standard errors in parentheses are clustered at the community level. \* $p < 0.1$ , \*\* $p < 0.05$ , \*\*\* $p < 0.01$ .

## Footnotes

1 See for example Good Morning America (2020), CBS News (2021) and Time (2020).

2 An analogy is to a game of Russian Roulette. The more live bullets in the chamber, the greater the risk of dying (case counts). If one knows precisely which chamber has the live bullet, then one can manage their risk by deciding in each round whether to play or pass (uncertainty). The more times one plays, the greater the risk of dying (contact).

3 Temperature and symptom screening have limited effectiveness, since some infected individuals are asymptomatic.

4 Protective equipment may reduce this risk but does not eliminate it, and in resource-poor settings this is often a moot point as high-quality protection is generally not widely available. While the situation is vastly improved in high-income countries, in the early months of the pandemic demand often exceeded supply causing health workers to resort to unorthodox strategies (Ankel, 2020).

5 In the extreme, a health worker might even consider leaving the profession (Delaney et al., 2021).

6 The data come from a larger research project that will be described later.

7 This is not a novel prescription: grocery workers in the US, for example, received additional hazard pay during the Covid-19 pandemic (Kinder and Stateler, 2021). Such payment adjustments are, in fact, not unusual in the health sector. Health workers, for example, are compensated for taking on more difficult assignments such as rural postings.

8 The states are Akwa Ibom, Gombe, Jigawa, and Kano. Nigeria has 36 states and one Federal Capital Territory.

9 In practice a research assistant waited somewhere they could observe the entrance to the treatment room and approached patients after they emerged. The goal was to interview five patients in each facility during each visit.

10 Pregnancy was self-reported.

11 Values between 0.41 and 0.6 are considered moderate agreement, values between 0.61-0.8 are considered substantial agreement, and values between 0.81 and 1 are considered perfect agreement (Viera et al., 2005).

12 To identify postpartum infections we asked women whether they experienced a foul-smelling vaginal discharge or pus within the first six weeks after delivery. For bleeding, we asked women if they lost a lot of blood during labor or delivery, or within 1-2 days after. ‘A lot of blood’ is obviously open to interpretation and so one might expect that the reported rate will probably be higher than if it were clinically measured.

13 We did not ask about postpartum infections because we were not convinced that women would be able to reliably recall this after so long.

14 Health worker characteristics are described in Table A.4.

15 Though not strictly comparable, this is not that different from results obtained in other settings (Geldsetzer, 2020) though one might hope that health workers would have more accurate beliefs.

16 Figure A.1 shows that health worker beliefs about Covid-19 mortality are positively correlated with their level of concern about getting infected. They were asked how worried they were about getting infected on a scale from 0-10.

17 College-educated health workers include doctors (who are awarded a Bachelor of Medicine, Bachelor of Surgery on graduation), nurses with a Bachelor of Science in Nursing, and other health workers with a Bachelor’s degree in another discipline, e.g., pharmacy or public health, or a Higher National Diploma which is the equivalent.

18 There is no evidence of heterogeneity across intervention groups. I cannot reject the null for any of the outcomes.

19 Knowledge of this setting suggests that it would be extremely unusual for providers to be this solicitous of patients.

20 For the unscheduled visits, having no patients in the waiting area was not uncommon (28% of observations are zero) so to avoid dropping these observations I add one before log transforming. The results are unchanged if the zeros are not included.

21 Difficulty breathing was a close third but only nine patients (total) complained of difficulty breathing.

22 The aggregate number of deliveries in each month is shown in Table A.10.

23 I cannot perform the same test for 2019 because there were only a few births over the relevant time period (recall that women were pregnant at enrollment in 2019). In the four months before/after April 2019 there were only 334 total deliveries.

24 In discussing the study findings with health workers in the US, they related stories of avoiding chest examinations of patients suspected to have Covid-19, and active avoidance, on the hospital floor, of patients confirmed to have Covid-19 leading to delays in providing them with care.

## Notes

This study was supported by a grant from the Eunice Kennedy Shriver National Institute for Child Health and Human Development (R01HD090231). The funder played no role in the design and conduct of the study; collection, management, analysis, and interpretation of the data; preparation, review, or approval of the manuscript; and decision to submit the manuscript for publication. Excellent research assistance was provided by Rebecca De Guttry.

### Competing Interest Statement

The authors have declared no competing interest.

### Funding Statement

This study was supported by a grant from the Eunice Kennedy Shriver National Institute for Child Health and Human Development (R01HD090231).

### Author Declarations

Ethical approval for the research project was granted by the RAND Human Subjects Protection Committee and the Ethics Committee of Aminu Kano Teaching Hospital, Kano.

